# Concurrent validity, test-retest reliability, and normative properties of the Ignite app: a cognitive assessment for frontotemporal dementia

**DOI:** 10.1101/2024.05.06.24306341

**Authors:** Rhian S. Convery, Kerala Adams-Carr, Jennifer M. Nicholas, Katrina M. Moore, Sophie Goldsmith, Martina Bocchetta, Lucy L. Russell, Jonathan D. Rohrer

## Abstract

Digital biomarkers can provide frequent, real-time monitoring of health-related behaviour and could play an important role in the assessment of cognition in frontotemporal dementia (FTD). However, the validity and reliability of digital biomarkers as measures of cognitive function must first be determined. The Ignite computerised cognitive app contains several iPad-based measures of executive function, social cognition, and other domains known to be affected in FTD. Here we describe the normative properties of the Ignite tests, evaluate the associations with gold-standard neuropsychology tests, and investigate test-retest reliability through two healthy controls studies. Over 2,000 cognitively normal adults aged 20-80 years (mean=55.2, standard deviation=15.8) were recruited to complete the Ignite app through a remote data collection study. Significant associations were found between age and performance on several Ignite measures of processing speed (*r*=0.42 to 0.56, *p*<0.001) and executive function (*r*=0.43 to 0.62, *p*<0.001), suggesting the tests are sensitive to cognitive decline observed in normal ageing. A separate cohort of 98 healthy controls were recruited to an observational study (mean age=51.2 years, standard deviation=17.3), completing Ignite at two timepoints (7 days apart), a gold-standard pen and paper neuropsychology battery of corresponding tests, and a user experience questionnaire (10-items). The Ignite tests demonstrated moderate to excellent test-retest reliability (ICCs=0.54 to 0.92) and significantly correlated with their pen and paper counterparts (*r*=0.25 to 0.72, *p*<0.05). The majority of participants (>90%) also rated the app favourably, stating it was enjoyable and easy to complete unsupervised. These findings suggest the Ignite tests are valid measures of cognitive processes, capture a stable picture of performance over time and are well accepted in healthy controls, speaking to the feasibility of administering the app remotely. Therefore, the results have important implications for the utility of Ignite as a cognitive endpoint in upcoming FTD clinical trials.

## Introduction

Frontotemporal dementia (FTD) is a clinically, genetically, and pathologically diverse neurodegenerative disorder. Clinically, FTD is characterised by behavioural change (behavioural variant FTD) or language impairment (primary progressive aphasia) and can overlap with amyotrophic lateral sclerosis and atypical parkinsonian disorders. The majority of FTD cases are sporadic, however, in approximately one third of individuals symptoms are caused by an autosomal dominant genetic mutation in the *C9orf72*, *MAPT*, or *GRN* genes (1,2). Evaluating first-degree relatives of individuals with genetic FTD provides an insight into the presymptomatic stage of the disease, where individuals do not have symptoms but have a 50% chance of carrying one of the mutations.

There are currently no disease modifying therapies available for FTD, however, clinical trials that target pathogenic mutations are underway. These therapies will likely be the most effective when administered early in the disease course (3). However, sensitive and validated biomarkers of disease onset and progression in the presymptomatic phase of FTD are lacking, particularly cognitive ones. Traditional pen and paper neuropsychology tasks are laborious, require participants to break from their normal routine, and lack reliability and consistency in administration and scoring. In addition, tasks appear to lack sensitivity to cognitive change in FTD prior to symptom onset (4,5). The ubiquitous use of technology may provide a solution for improving the current standard of cognitive assessments. Digital cognitive assessments can reduce inter-rater variability, time, and associated costs of testing, and can be used as home monitoring tools, reducing patient burden and capturing cognition in a participant’s natural environment. Furthermore, such assessments allow for frequent testing, thus ensuring studies have more detailed datasets, and potentially yielding more sensitive measures of cognition. As such, several digital cognitive assessments have been developed for the early detection of mild cognitive impairment (MCI) both for screening tools and clinical endpoints in Alzheimer’s disease (AD) trials (6–11). However, there are few digital cognitive assessments available that are specifically designed to measure cognitive impairment in FTD.

Ignite is a cognitive assessment app for the iPad, designed for FTD observational research and clinical trials. The app includes 12 unique tests measuring information processing speed, executive function, social cognition, semantic knowledge, arithmetic, and visuospatial skills (12). Tests were included to tap into domains known to be affected in FTD, particularly presymptomatically, with the goal of improving sensitivity to detect cognitive impairment (13–16). Ignite is a self-assessment and is mainly comprised of computerised versions of standard neuropsychology tasks that can be completed in under 30 minutes. Before being tested in clinical cohorts to assess impairment, novel cognitive assessments need to provide appropriate validity and reliability estimates, including the development of new normative properties (17,18). Furthermore, demonstrating the feasibility of administering computerised cognitive assessments is equally important to ensure digital assessments are well accepted in the population. Here we describe the normative properties and construct validity of the Ignite tests in a study of over 2,000 healthy controls (Study 1) and report concurrent validity with gold-standard neuropsychometry, test-retest reliability estimates, and feasibility data in a separate study of 98 healthy controls (Study 2).

## Methods

### STUDY 1

#### Participants

Participants were recruited to complete Ignite remotely via media advertisements and were eligible to take part if they were healthy controls (i.e. did not have a significant neurological or psychiatric disorder), were aged between 20-80, owned an Apple iPad (any model), and were able to understand and comply with instructions in English. Participants were encouraged to read the study information sheet on the first page of the app to ensure they met the criteria.

#### Procedure

Instructions were provided in the study advertisement describing how to download the Ignite app from the App Store onto participant’s personal devices. After reading the information sheet in the app and completing the consent form, participants were then presented with a short form requiring them to enter basic demographic information, including age (years), education (years), sex (M/F), country of residence, and the first three letters of their city of birth (to help with the identification of duplicate attempts at the tests). No personally identifiable information was collected. Participants then completed 12 separate cognitive tasks in a predetermined order: Think Back Level 1 and Level 2, Sum Up, Colour Mix Levels 1-4, Face Match, Mind Reading, Swipe Out, Card Sort, Line Judge, Balloon Fair, Time Tap, Path Finder Levels 1 and 2, and Picture Pair (Table 1, Figure 1). Detailed instructions were presented at the beginning of each test accompanied by example videos demonstrating how the task should be completed. No feedback was provided to participants on their performance, and on the final page of the assessment participants were required to select “Upload data”, which sent the results to a secure server.

**Figure 1:** Tasks in the Ignite battery: please contact the corresponding author to request access to the Ignite test images.

**Table 1:** Ignite tests with corresponding gold-standard pen and paper equivalents, the cognitive (subdomain) measured, a description of the task, the maximum number of possible trials, and the time allowed to complete each test (in seconds). D-KEFS = Delis-Kaplan Executive Function System.

| **Ignite test** | **Gold-standard test** | **Cognitive domain**<br>**(subdomain)** | **Description** | **Number of**<br>**trials** | **Time to**<br>**complete**<br>**(s)** |
| --- | --- | --- | --- | --- | --- |
| Think Back (Level 1) | N-back (1 back) | Processing speed | Participants must indicate whether a shape presented exactly matches (including colour) the shape that preceded it. | 72 | 60 |
| Think Back (Level 2) | N-back (2 back) | Executive function<br>(Working memory) | Participants must indicate whether a shape presented exactly matches (including colour) the shape that came two before it. | 72 | 60 |
| Sum Up | Graded Difficulty<br>Arithmetic Test | Arithmetic | Participants are required to select the correct answer (from four options) to different calculations including additions, subtractions, multiplications, and divisions. | 48 | 60 |
| Colour Mix (Level 1) | D-KEFS - Color<br>Naming | Processing speed | Participants are required to tap the name of a colour that matches the colour of a circle. | 50 | 30 |
| Colour Mix (Level 2) | D-KEFS - Word<br>Reading | Processing speed | Participants are required to match the name of a colour that matches a colour word. | 50 | 30 |
| Colour Mix (Level 3) | D-KEFS – Color-Word Inhibition | Executive function<br>(Inhibitory control) | Participants are presented with a colour word written in different coloured ink. They are required to choose the name of the colour that matches the colour of the ink. | 50 | 60 |
| Colour Mix (Level 4) | D-KEFS -<br>Inhibition/Switching | Executive function<br>(Inhibitory<br>control/Set-shifting) | Participants are presented with a colour word written in different coloured ink. They are required to complete the task as in Level 3, unless a black border appears around the word in which case, they should match the written word rather than the ink colour. | 50 | 60 |
| Face Match | Ekman 60 Faces<br>Test | Social cognition | Participants are required to tap the faces displaying a target emotion that match a word written at the top of the screen (e.g., Happy, Surprise, Anger, Fear, Sadness, Disgust). | 30 | 60 |
| Mind Reading | Reading the Mind in<br>the Eyes task | Social cognition | Participants are required to tap the pair of eyes (from four options) that match a target emotion written at the top of the screen (e.g., reflective, contemplative). | 20 | 90 |
| Swipe Out | Eriksen Flanker task | Executive function<br>(Inhibitory control) | Participants are presented with arrows pointing in a given direction. They are required to swipe on the screen, in the direction of the central arrow. Flanking arrows are either facing in the same (congruent) or different (incongruent) direction. | 40 | 60 |
| Card Sort | Wisconsin Card Sorting Task | Executive function (Set-shifting) | Participants are presented with four cards, one in each corner of the screen, and one card in the middle. They are required to match the card in the middle to one of the four cards according to a secret rule (e.g., Colour, Shape or Number). | 48 | 90 |
| Line Judge | Benton Judgement of Line Orientation | Visuospatial skills | Participants are required to select two lines that match with the orientation of two target lines presented above. | 30 | 90 |
| Balloon Fair | Iowa Gambling Task | Executive function (Decision making) | Participants are required to pump a balloon to win money. The more the balloon is inflated the more money there is available to collect, however, if the balloon is pumped too much it can burst. | 50 | 90 |
| Time Tap | - | Executive function (Cognitive timing) | Participants are required to tap in time with a circle that pulses at regular intervals. After 30s the circle disappears, and the participant must continue tapping at the same tempo. | 40 | 60 |
| Path Finder (Level 1) | Trail Making Test Part A | Processing speed | Participants are required to tap from one number to the next in sequential order. | 19 | 90 |
| Path Finder (Level 2) | Trail Making Test Part B | Executive function (Set-shifting) | Participants are required to alternate between tapping from numbers to letters in sequence (i.e., 1, A, 2, B). | 19 | 90 |
| Picture Pair | Modified Camel and Cactus Test | Semantic Knowledge | Participants are required to select an image (from four options) that best matches with a target image. | 25 | 120 |

#### Data pre-processing

Data from each participant were collated and analysed in Stata/MP (version 16.1). Participants were grouped into six age groups (20-29, 30-39, 40-49, 50-59, 60-69, 70-80 years) and four education groups (0-9, 10-12, 13-16, and ≥17 years). These education groupings were chosen to reflect standard levels of education in the United Kingdom. For each test in the assessment, the number of trials completed per person were calculated and averaged across each age group. For each test in the assessment, participants were excluded if the number of trials they had completed was more than three standard deviations below the mean for their age group. This criterion was applied to ensure there were a sufficient number of trials to analyse for each test and to remove participants that had not properly attempted the task.

#### Outcome measures

For several tests, including Think Back, Colour Mix, Mind Reading, Face Match, Sum Up, Picture Pair, and Line Judge, three measures of speed (average reaction time across trials), accuracy (total correct), and a speed-accuracy trade-off score (SAT = total correct/average reaction time) were calculated for each test. Average reaction time was also calculated for the Swipe Out task along with the Flanker effect measure (average reaction time of incongruent trials - average reaction time of congruent trials). The total completion time (in seconds) was calculated for Path Finder Levels 1 and 2. The total number of correct categories achieved was the measure of interest for the Card Sort task, and the total amount of money won was calculated for the Balloon Fair test. Clock variance and absolute drift were computed from an autoregressive timing model for the Time Tap task (19), where clock variance represents a cognitive representation of time or “the internal clock”, and absolute drift is the difference in the response interval from the first and last tap (see Supplementary Table 1 for a detailed description of outcome measure calculation).

#### Statistical analysis

### Demographic associations

To examine the isolated effects of demographic predictors on performance, the partial correlations of age and education with each of the Ignite outcome measures were calculated, controlling for the remaining demographic variables (i.e., sex and age/education). Pearson’s or Spearman’s (if data was non-normal) partial correlations were performed in RStudio Team (2021). Linear regressions were used to analyse sex differences with males as the reference group, and age and education included as covariates in the model. For outcomes that were not normally distributed, bootstrapping with 2000 replications was used to calculate 95% confidence intervals. For the Card Sort task, as the outcome measure (number of correct categories) is categorical, a logistic regression was used to investigate if age, education, and sex predicted task performance. All regression analyses were performed in Stata/MP (version 16.1).

### Normative properties

To produce normative scores by age deciles, education groups, and sex, adjusted means were output from linear regression estimates for each Ignite outcome measure. To generate a *Z*-score calculator from the normative data, multiple linear regressions were conducted for each Ignite outcome measure that adjusted for the demographic predictors of age, sex, and years in education, concurrently, individually and without covariates, resulting in five different models per measure. *Z*-scores were then estimated by subtracting raw scores from the predicted mean(s) and then dividing this difference score by the standard deviation of the residuals (root mean squared error term). To ensure *Z*-scores were interpretable, outcome measures that were not normally distributed were transformed prior to analysis (see Supplementary Table 2).

### Construct validity

To understand construct validity, an exploratory factor analysis was performed. An iterated principal factor method was used to analyse a correlation matrix of Ignite outcome measures. Only continuous variables were included, excluding the Card Sort task from this analysis. To select the optimal number of factors, those with an eigenvalue greater than 1 were retained, resulting in a five-factor solution. Factor loadings were interpreted using the oblique promax rotation method and a minimum criterion of a primary loading factor of 0.3 or above was applied to each outcome measure (20).

### STUDY 2

#### Participants

Participants were recruited to an observational study held at University College London (UCL). All participants gave fully informed consent at the beginning of the research visit. The same inclusion and exclusion criteria used in Study 1 were applied, with the exception that participants did not need to own an Apple iPad, and an additional requirement that they must not have completed the Ignite app before (i.e., as part of Study 1). Healthy controls were recruited through the online platform Join Dementia Research (www.joindementiaresearch.nihr.ac.uk). Only individuals that met the study criteria received the details, and then these participants could choose to register to take part in the study.

#### Procedure

Participants attended two 1-hour research visits at UCL conducted two weeks apart. All participants completed the Ignite app and a neuropsychology battery containing gold-standard pen and paper versions of the Ignite tests. Participants were randomised 1:1 into two conditions, completing either the Ignite app or the neuropsychology battery at the first research visit. The pen and paper neuropsychology battery was administered in a quiet testing room and included 11 different tests (see Table 1). After the first research visit, participants were given a study iPad, with the Ignite app downloaded, and were asked to complete the assessment 1-week later. Therefore, participants completed Ignite at two timepoints (7 days apart), once remotely, and once during one of the research visits. At the end of the study, participants were invited to complete the Ignite User Experience questionnaire, via email link to the Lime Survey platform (version 2.28.34). The survey included 10 statements, concerning attitudes and experiences of completing Ignite, rated on a 5-point Likert scale (ranging from “Strongly disagree” to “Strongly agree”). To reduce response bias, statements were randomised so that 50% were of a positive attitude and 50% were negative.

#### Statistical analysis

### Concurrent validity

A correlation analysis was conducted to determine the relationship between Ignite tests and the standard pen and paper neuropsychology tasks. Pearson correlations were computed for normal measures and Spearman correlations were used for outcome measures with a non-normal distribution. A chi-squared test was conducted to assess the relationship between scores on the Card Sort task and the Wisconsin Card Sorting Task. The most comparable outcome measures were chosen to compare performance on Ignite tests with the neuropsychology battery. Where direct equivalents were not available, Ignite tests were correlated with other pen and paper tasks hypothesised to measure the same cognitive domains.

### Test-retest reliability

A two-way mixed effects model for each Ignite outcome measures across the two timepoints was used to calculate consistency of agreement intraclass correlation coefficients (CA-ICC) (21). Consistency of agreement ICCs were selected to allow for a difference in performance upon repeated assessment (e.g. learning effects) (22). Established cut-off criteria for CA-ICC values were used to determine the level of reliability for each outcome measure where: poor ≤ 0.50, moderate=0.50-0.75, good=0.75-0.90, excellent ≥ 0.90 (22,23). The mixed model was also used to calculate the mean difference between scores and the two time points and the 95% upper and lower limits of agreement for the mean difference. Non-normally distributed data were transformed prior to analysis. Bland-Altman plots were constructed to demonstrate agreement between Ignite scores, and back-transformed for measures that were not normally distributed.

### User experience questionnaire

The percentage of responses were calculated for each rating on the Likert scale. “Strongly Agree” and “Agree” responses were collated into one single measure of “Agreement” and “Strongly Disagree” and “Disagree” responses into one measure of “Disagreement” to improve the interpretability and provide an overall picture of the group attitude per question. To investigate potential differences in user experience by age, participants were split into two groups of younger (age <59 years) and older (age ≥ 60) adults, and chi-squared tests were used to assess differences in rating for each question.

## Results

### STUDY 1

#### Participant characteristics

A total of 2,043 people completed the Ignite app. There were 9 different countries represented in the dataset, with 95.3% of participants residing in the United Kingdom and 4.3% from the United States (US). The remaining 0.4% (*N*=8) of participants were from countries where English was not the predominant language and were therefore excluded to limit the effects of language comprehension on task performance. Analysis of the data revealed 31 participants with identical demographic information to at least one other participant. These individuals were also excluded based on the possibility these could be duplicate participants, resulting in 39 healthy controls in total being excluded prior to analysis. Therefore, 2,004 participants were included in this study; see Table 2 for demographic information. Female participants accounted for 67.4% of the normative sample. The mean (standard deviation) age of the population was 55.2 (15.8), and the number of years in education was 16.1 (4.2).

**Table 2:**
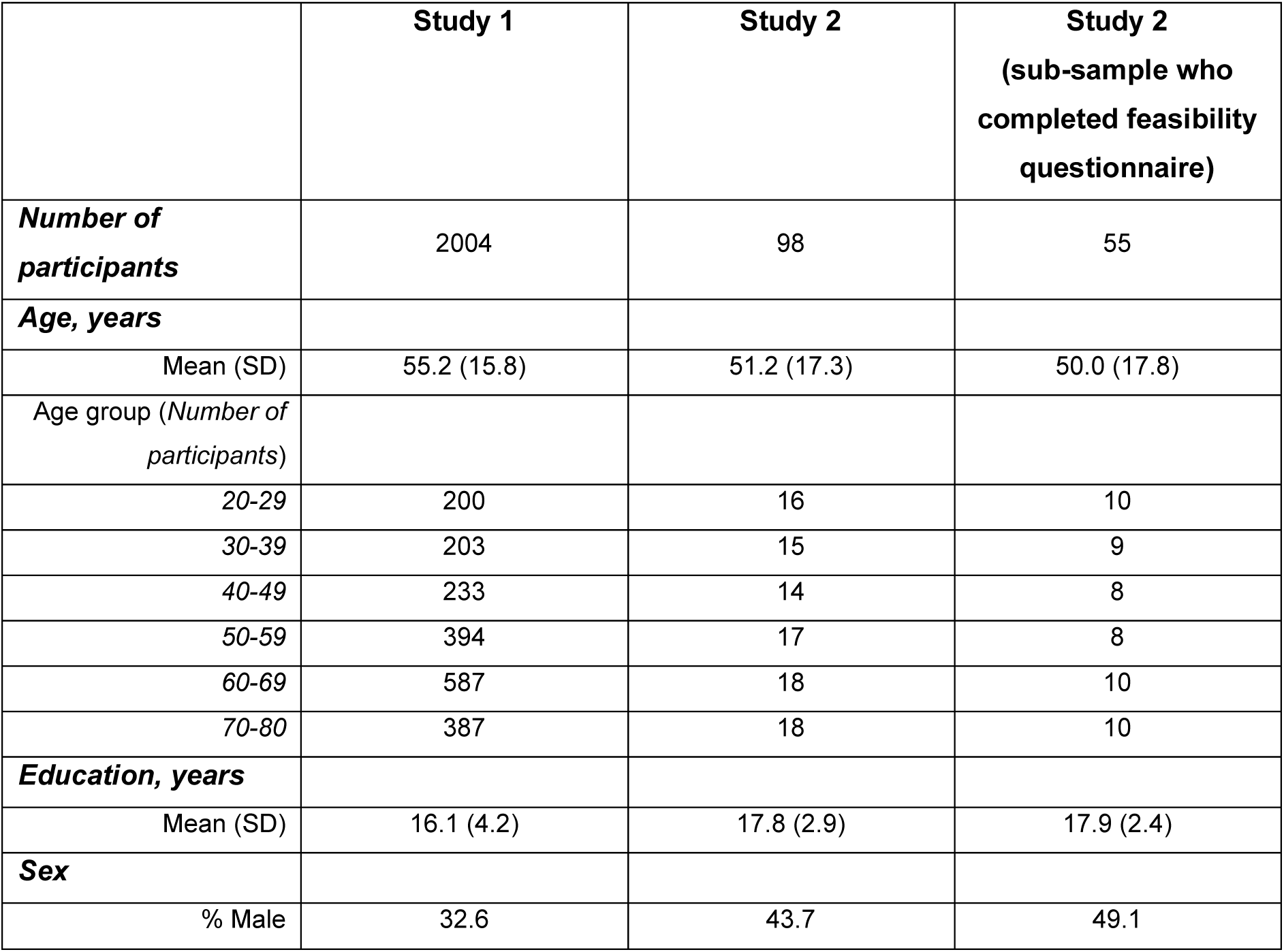
Demographic characteristics of the healthy control participants in each study. SD = standard deviation.

#### Demographic associations

Significant associations between age and performance were seen for 38 out of the 43 Ignite outcome measures (*p*<0.01). A positive correlation between age and average reaction time was observed across the tests (*r*=0.24 to 0.62, *p*<0.001), indicating a slowing of responses with age, accompanied by a decrease in accuracy in the total number of correct trials (*r*=-0.12 to -0.57, *p*<0.001) and SAT scores (*r*=-0.25 to -0.58, *p*<0.001), see Figure 2. A decline in performance was also seen with age and the total money earned on the Balloon Fair task (*r*=-0.36, *p*<0.001), and the number of correct categories achieved in the Card Sort test (β=-0.06, *p*<0.001). Only the Sum Up task did not display significant correlations with age. Significant sex differences were observed with females performing better on Colour Mix Levels 1-3 (β=0.39 to 1.25, *p*<0.05), Path Finder Level 2 (β=-2.30, *p*<0.001), Face Match (β=0.97 to 1.25, *p*<0.001), Mind Reading (β=0.55, *p*<0.001), and Picture Pair (β=0.43, *p*<0.05). Male participants had significantly higher scores on Sum Up (β=1.92, *p*<0.001), Line Judge (β=1.52 , *p*<0.001), Think Back Level 2 (β=1.82, *p*<0.001), and Balloon Fair tasks (β=85.5, *p*<0.001). Significant associations between education and performance were seen on several Ignite tests, however, effect sizes were small (see Table 3).

**Figure 2:**
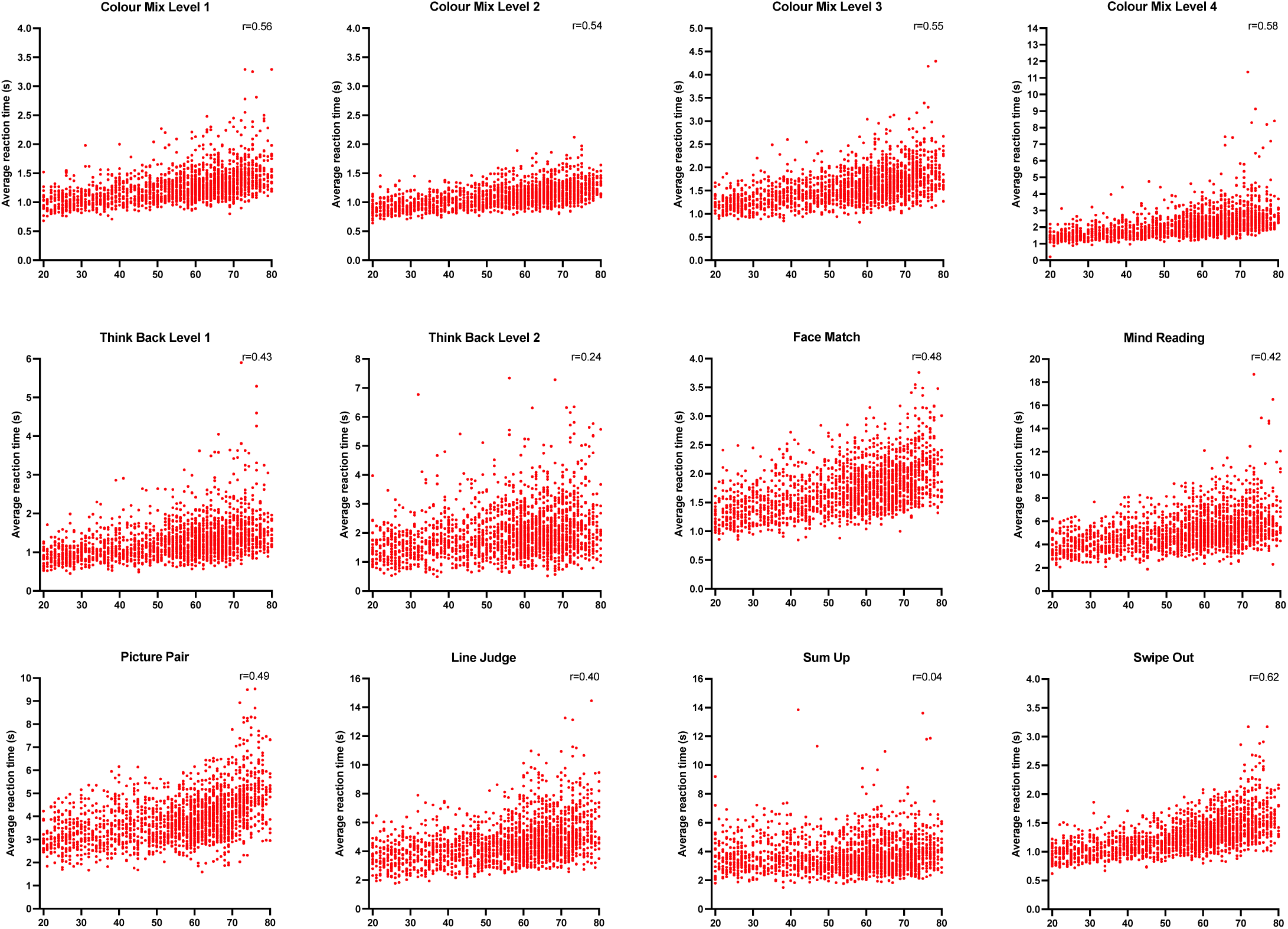
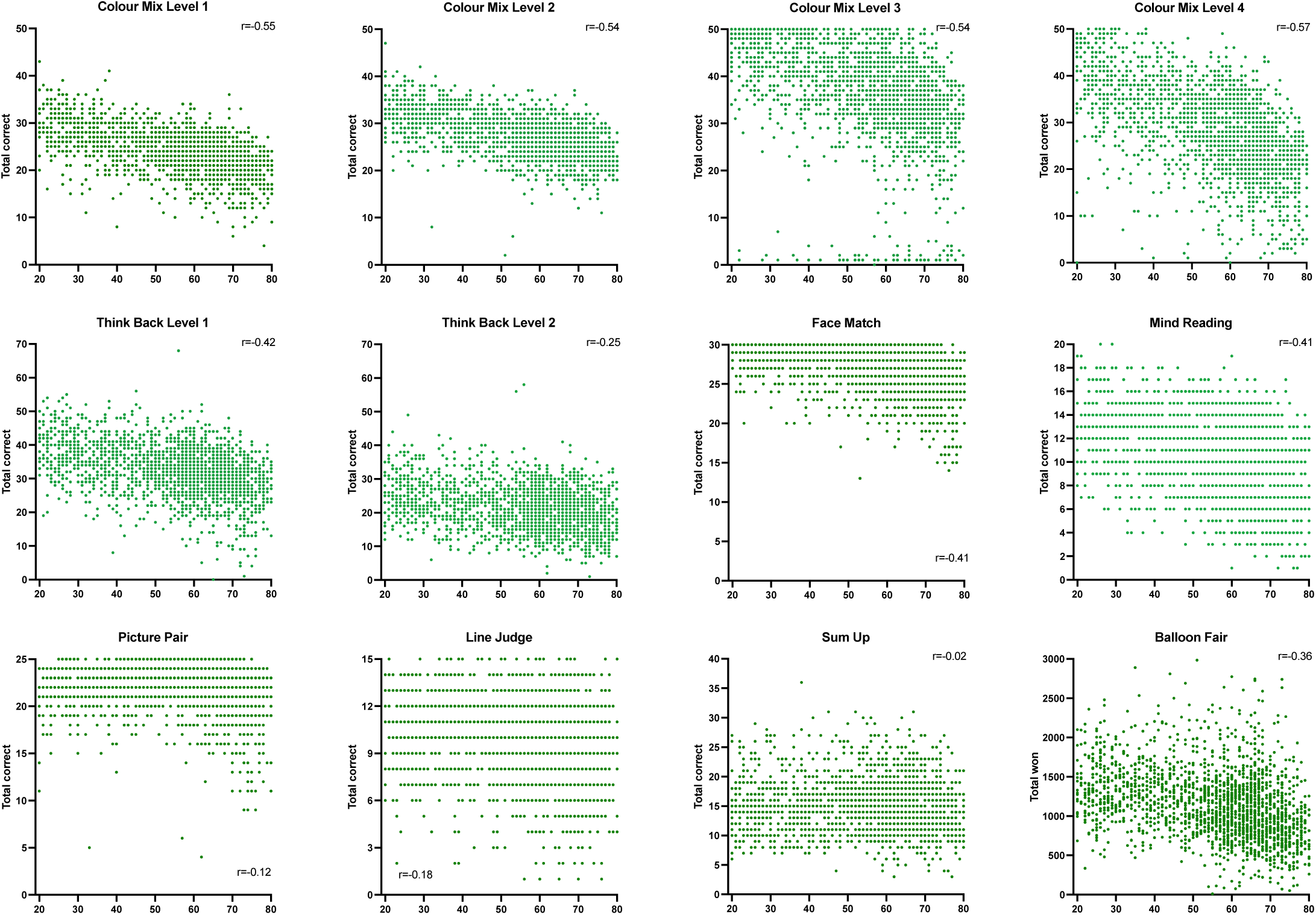
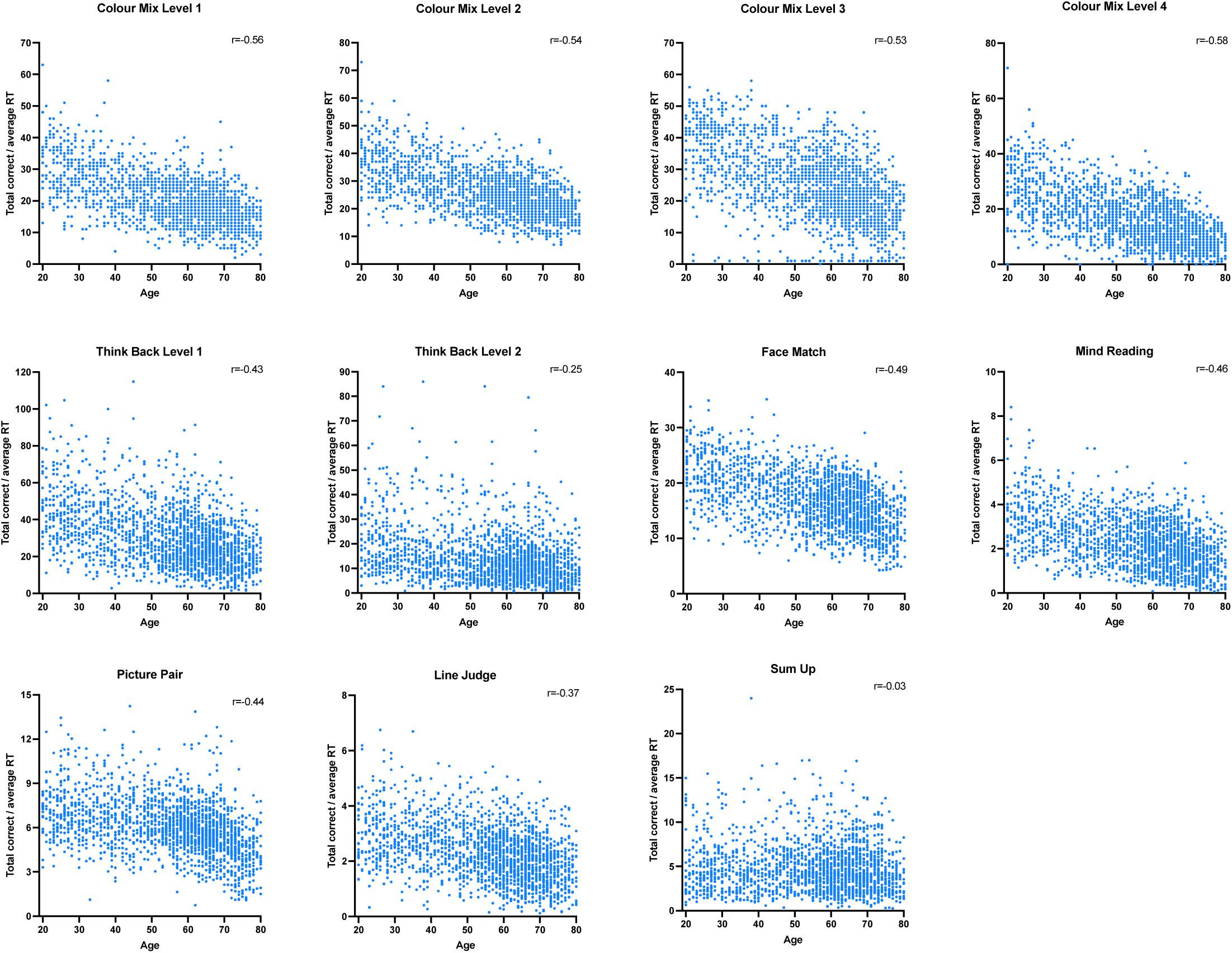
Scatterplots displaying the relationship between a) average reaction time (in seconds), b) total correct and c) SAT scores with age on the Ignite tests. r= partial correlation coefficient.

**Table 3:**
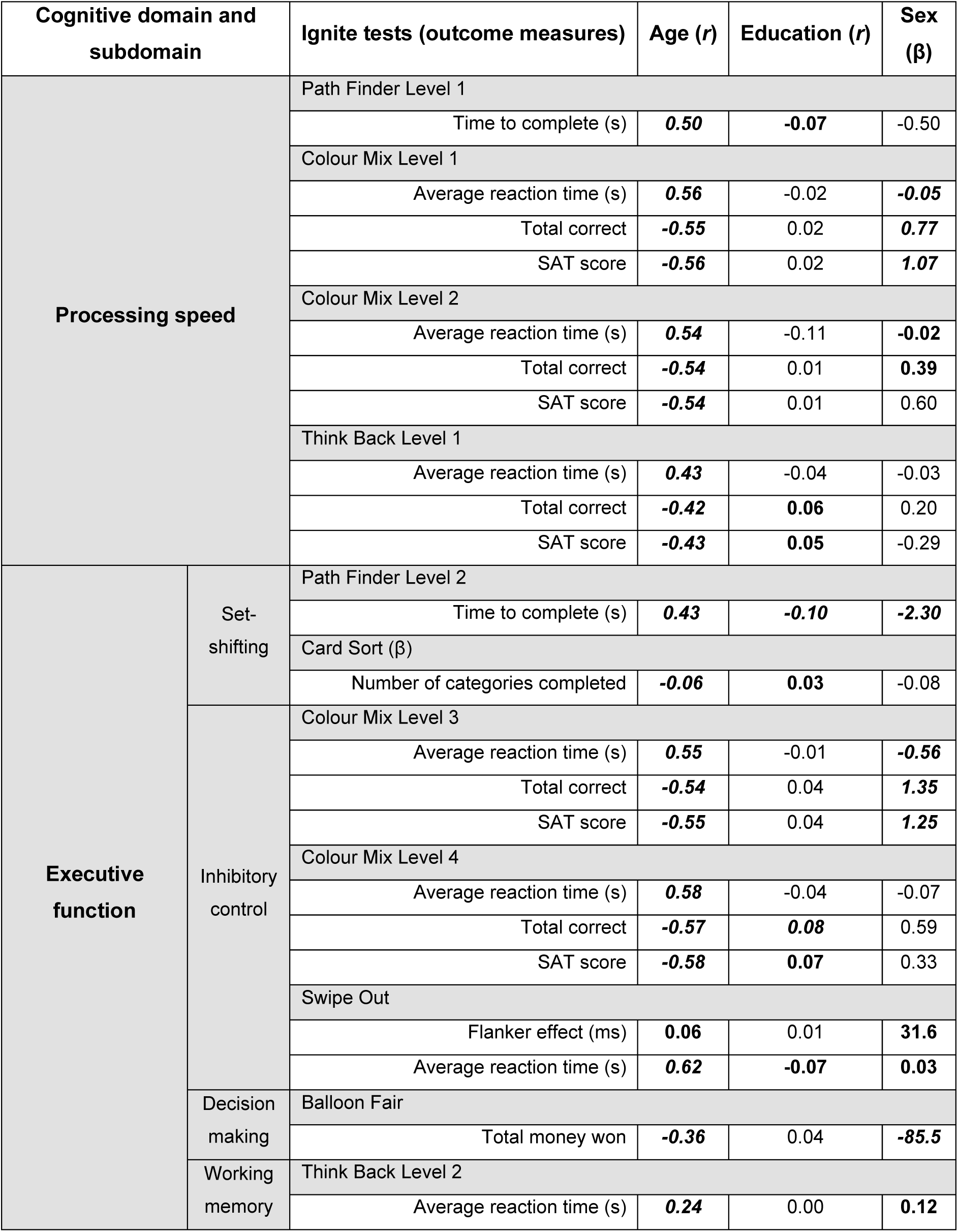
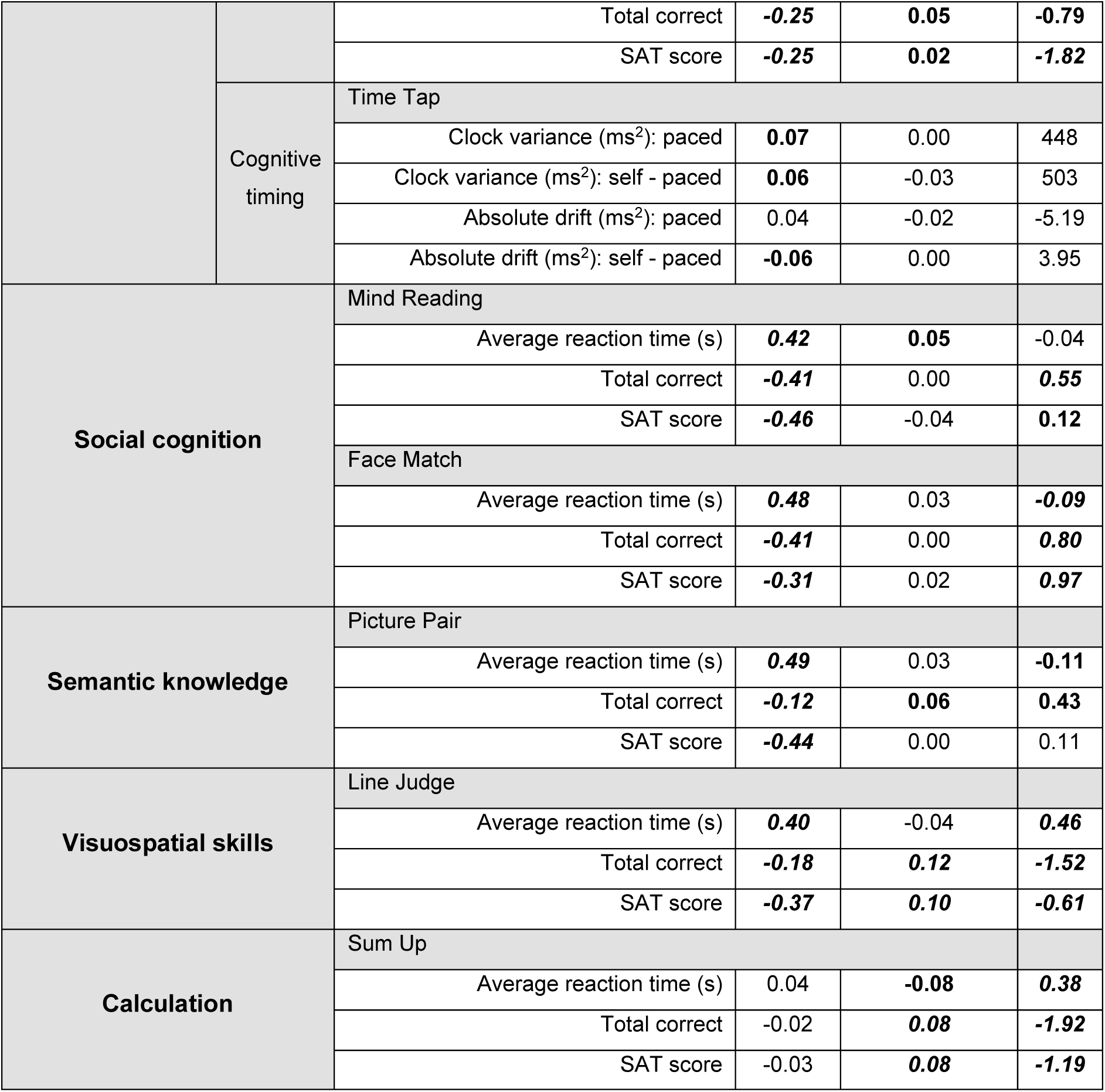
Partial correlations for age and education, and linear regression for sex for each Ignite outcome measure. *r*= correlation coefficient, β= linear regression coefficient with males as the reference group, SAT= speed-accuracy trade-off score. Bold and italicised values denote significance levels where *p*<0.001, and bold values indicate *p*<0.05.

**Figure 3:**
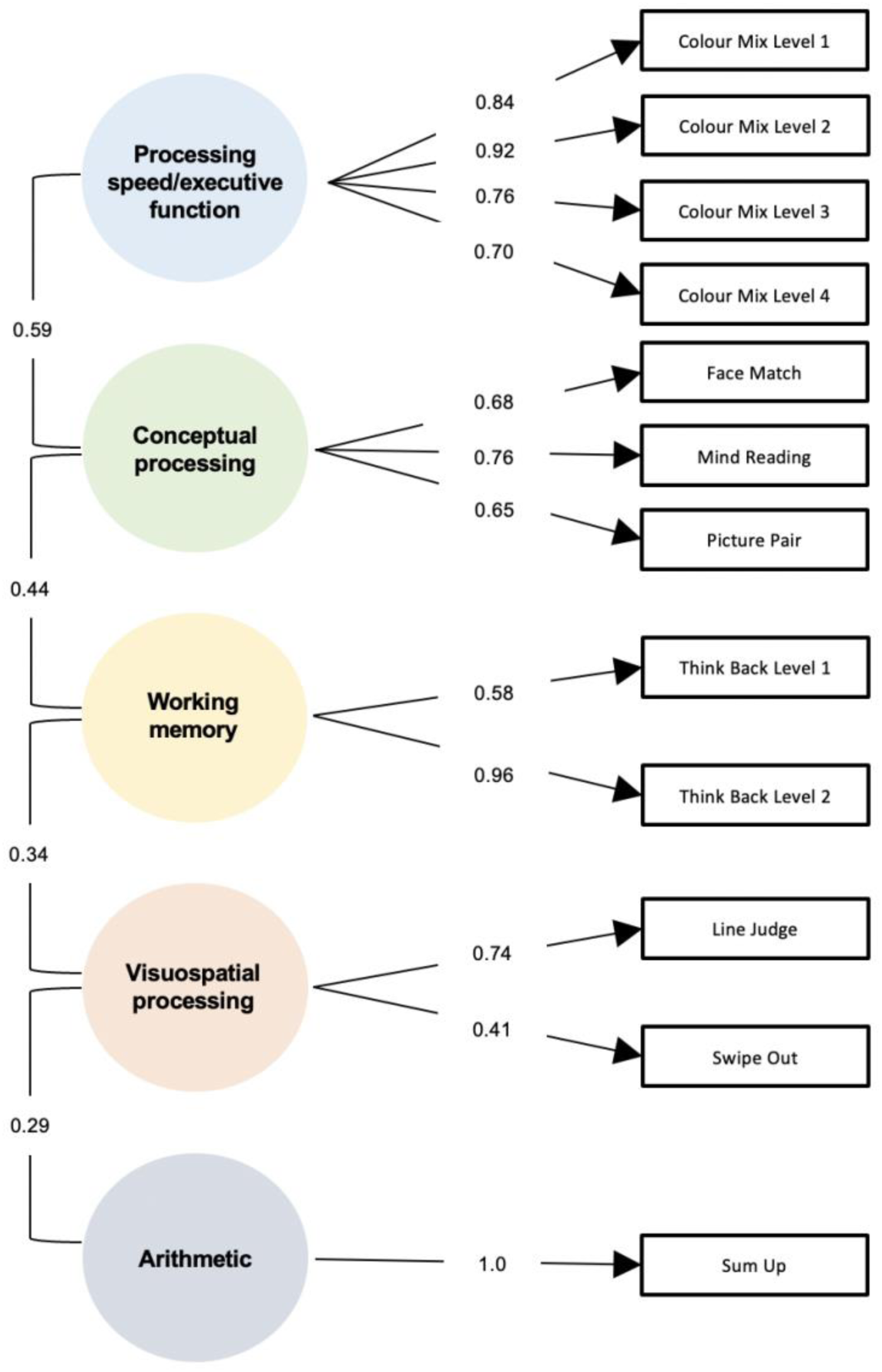
Diagram representing the factor structure of the Ignite tests, with circles labelled according to the cognitive domain each factor represents. The values denote the highest factor loading for an outcome measure in each test. Between factor correlations are also shown. Conceptual processing=social/semantic processing.

**Figure 3:**
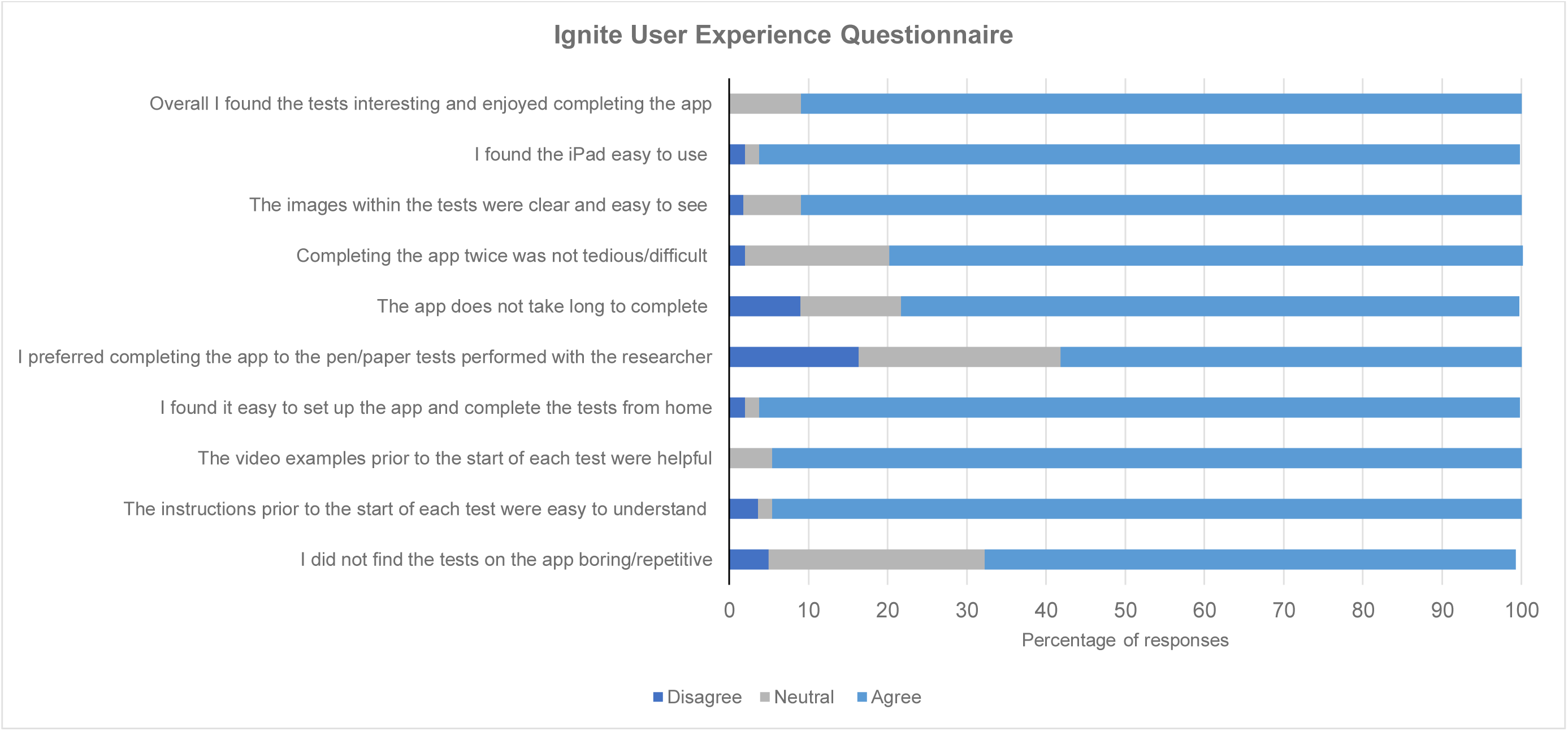
Stacked bar chart displaying the percentage of healthy controls that agree, disagree, or feel neutral to each statement in the Ignite User Experience questionnaire. Statements of a negative attitude were inversed for interpretability.

#### Normative data

Equations for calculating the *Z*-scores for each outcome measure were used to generate a normative calculator for raw scores. Percentile ranks were also calculated from the normal distribution of each *Z-* score. The calculator was constructed to provide five corresponding estimated *Z*-scores, per Ignite outcome measure, based on predictions from each linear regression model. Therefore, an individual’s raw Ignite scores and demographic information can be input into the spreadsheet, and *Z*-scores and percentile ranks are subsequently generated for each adjustment. The normative calculator is now available on the Genetic Frontotemporal dementia Initiative (GENFI) website (www.genfi.org/ignite).

Age- and education-grouped normative values, as well as differences in sex, for each Ignite outcome measure are reported in Supplementary Table 3.

#### Construct validity

The five-factor solution from the factor analysis explained 87.9% of the variance in the data. All outcome measures from individual tests (i.e., average reaction time, total correct, and SAT scores) loaded on the same factors. Colour Mix Levels 1, 2, 3, and 4 were contained in Factor 1, whilst Face Match, Mind Reading, and Picture Pair loaded onto Factor 2 (see Table 4). Both Think Back Levels 1 and 2 loaded with Factor 3. Outcome measures from Line Judge and Swipe Out loaded together in Factor 4, and finally Sum Up outcome measures were contained within Factor 5. Cross-loading was minimal but was observed for Think Back Level 1 and Swipe Out (with Factor 1) and Colour Mix tasks (with Factor 4). Factors were subsequently grouped with the following labels: (1) Processing speed/executive function, (2) Social/semantic processing, (3) Working memory, (4) Visuospatial processing, and (5) Arithmetic (see **Error! Reference source not found.**). Path Finder Levels 1 and 2 were just below the minimum criteria (>0.3) of the primary loading factor for Factor 1 (−0.295 and -0.285, respectively). In addition, Balloon Fair and Time Tap tasks did not group under a simple factor structure, loading equally across factors.

**Table 4:**
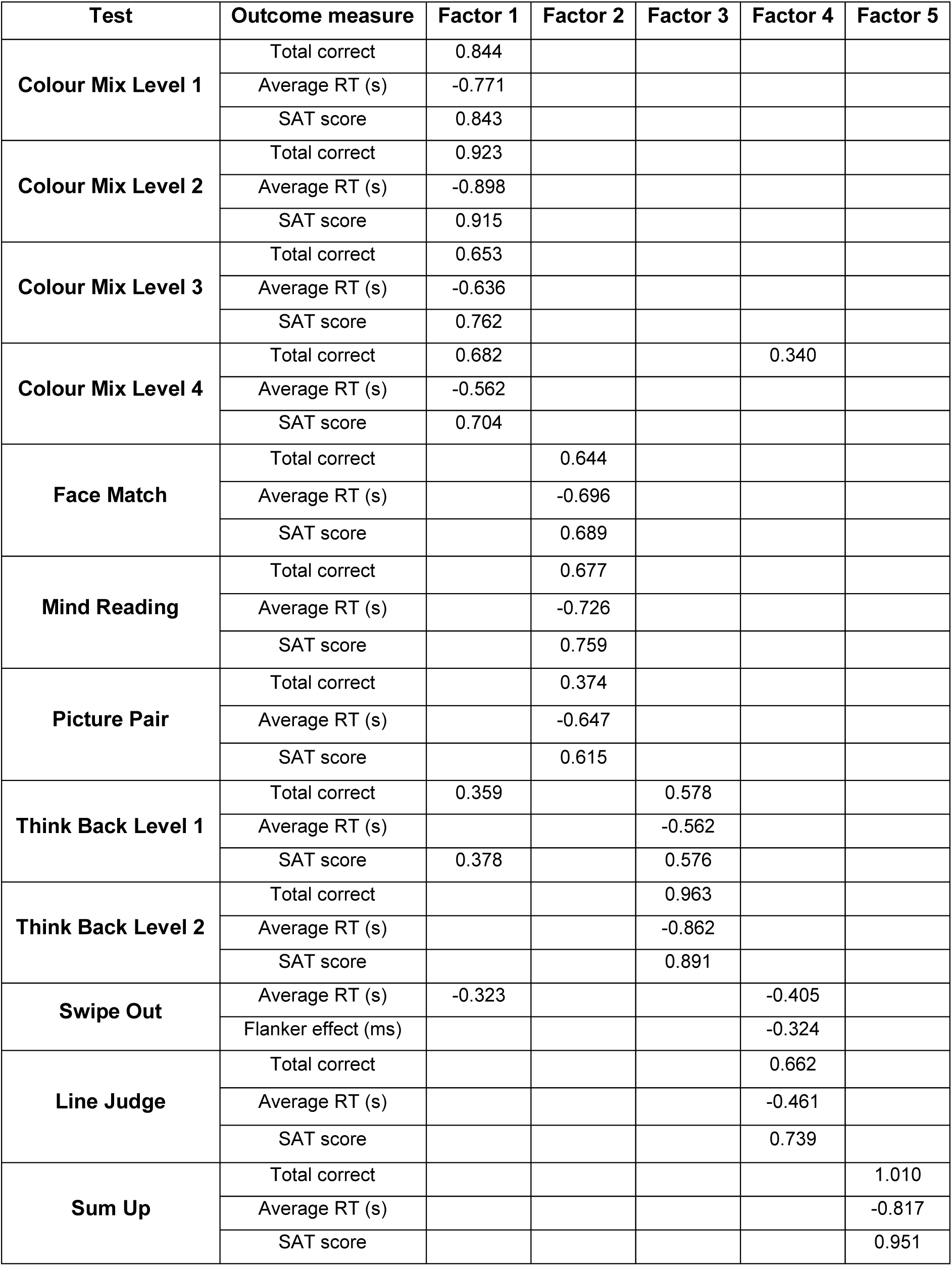
Rotated factor loadings for the five-factor model of Ignite outcome measures. Loadings greater than 0.3 are shown. RT=reaction time, SAT=speed accuracy trade-off.

### STUDY 2

#### Participant characteristics

A total of 98 healthy controls (43.7% male) with a mean (standard deviation) age of 51.2 (17.3) years, and number of years in education of 17.8 (2.9), were recruited (Table 2). A subset of participants (*N*=55) completed the Ignite User Experience questionnaire. This sub-sample had a mean (standard deviation) age of 50.0 (17.8) years and number of years in education of 17.9 (2.4), and 47.3% of the sample were male.

#### Concurrent validity

The majority of the Ignite tests significantly correlated with their pen and paper counterparts (*r*=0.25 to 0.72, *p*<0.05), and other neuropsychology tests measuring the same cognitive domains where direct comparisons were not available (*r*=0.31 to 0.73, *p*<0.05) (Table 5). Only the Face Match, Card Sort, and Time Tap tasks did not significantly correlate with corresponding neuropsychology tests.

**Table 5:** Correlations between Ignite and pen and paper neuropsychology tasks. RT=reaction time. D-KEFS = Delis-Kaplan Executive Function System.

| **Ignite test**<br>**(outcome measure)** | **Pen and paper test**<br>**(outcome measure)** | **Correlation**<br>**coefficient** | **p-value** |
| --- | --- | --- | --- |
| Path Finder Level 1 (completion time) | Trail Making Test Part A (completion time) | 0.55 | <0.001 |
| Path Finder Level 2 (completion time) | Trail Making Test Part B (completion time) | 0.52 | <0.001 |
| Colour Mix Level 1 (average RT) | D-KEFS - Color Naming (completion time) | 0.50 | <0.001 |
| Colour Mix Level 2 (average RT) | D-KEFS - Word Reading (completion time) | 0.25 | 0.013 |
| Colour Mix Level 3 (average RT) | D-KEFS - Color-Word Inhibition (completion time) | 0.71 | <0.001 |
| Picture Pair (total correct) | Modified Camel and Cactus Test (total correct) | 0.42 | <0.001 |
| Line Judge (total correct) | Benton Judgement of Line Orientation (total correct) | 0.58 | 0.000 |
| Face Match (total correct) | Ekman 60 Faces Test (total correct) | 0.17 | 0.098 |
| Mind Reading (total correct) | Reading the Mind in the Eyes task (total correct) | 0.38 | <0.001 |
| Sum Up (total correct) | Graded Difficulty Arithmetic Test (total correct) | 0.72 | <0.001 |
| Balloon Fair (total won) | Iowa Gambling Task (net total) | 0.31 | 0.004 |
| Swipe Out (average reaction time) | Eriksen Flanker Task (average reaction time) | 0.52 | <0.001 |
| Think Back Level 2 (total correct) | N-back (2 back) (total correct) | 0.46 | 0.002 |
| **Tests without standard equivalent** |  |  |  |
| Colour Mix Level 4 (average RT) | D-KEFS - Color-Word Inhibition (completion time) | 0.73 | <0.001 |
| Think Back Level 1 (average RT) | D-KEFS - Color Naming (completion time) | 0.44 | <0.001 |
| Think Back Level 1 (average RT) | Trail Making Test Part A (completion time) | 0.52 | <0.001 |
| Time Tap - Paced (clock variance) | D-KEFS - Color-Word Inhibition Test (completion time) | 0.13 | 0.211 |
| Time Tap - Paced (absolute drift) | Trail Making Test Part B (completion time) | 0.15 | 0.157 |
| Time Tap - Self-paced (clock variance) | D-KEFS - Color-Word Inhibition (completion time) | 0.08 | 0.471 |
| Time Tap - Self-paced (absolute drift) | Trail Making Test Part B (completion time) | 0.02 | 0.840 |

#### Test-retest reliability

With the exception of the Time Tap task (ICC’s = -0.12 to 0.12), the Ignite tests demonstrated moderate to excellent test-retest reliability estimates (ICC’s = 0.54 to 0.92) (Table 6). High levels of agreement were also observed in the majority of Ignite outcome measures as demonstrated by Bland-Altman plots (see Supplementary Figure 1).

**Table 6:** Test-retest reliability data for Ignite outcome measures. Data represents mean (standard deviation) scores on Ignite tests for Timepoint 1 and Timepoint 2 (7 days later) as well as the mean (standard deviation) for the difference between timepoints. Intraclass correlation coefficients measuring consistency of agreement (CA-ICC) between scores, the 95% confidence intervals (CI), and the upper and lower 95% limits of agreement are shown.

| Test Name | Outcome measure | Timepoint 1 | Timepoint 2 | Difference | CA-ICC | Lower CI | Upper CI | Lower agreement | Upper agreement |
| --- | --- | --- | --- | --- | --- | --- | --- | --- | --- |
| Sum Up | Total correct | 15.5 (5.06) | 17.1 (5.18) | 1.62 (2.32) | 0.90 | 0.85 | 0.93 | -3.01 | 6.26 |
|  | Average RT | 3.74 (1.29) | 3.34 (1.01) | -0.40 (0.67) | 0.89 | 0.84 | 0.93 | -0.38 | 0.18 |
|  | SAT score | 4.99 (3.10) | 5.99 (3.48) | 1.00 (1.36) | 0.88 | 0.83 | 0.92 | -0.39 | 0.81 |
| Colour Mix Level 1 | Total correct | 24.3 (5.05) | 25.9 (5.35) | 1.63 (3.19) | 0.81 | 0.73 | 0.87 | -4.75 | 8.01 |
|  | Average RT | 1.27 (0.32) | 1.18 (0.29) | -0.09 (0.22) | 0.82 | 0.74 | 0.88 | -0.15 | 0.27 |
|  | SAT score | 20.9 (8.29) | 23.9 (9.57) | 3.02 (5.20) | 0.82 | 0.74 | 0.88 | -0.83 | 1.46 |
| Colour Mix Level 2 | Total correct | 27.5 (4.50) | 28.5 (5.12) | 0.96 (2.70) | 0.84 | 0.77 | 0.89 | -4.44 | 6.36 |
|  | Average RT | 1.09 (0.19) | 1.06 (0.20) | -0.03 (0.12) | 0.84 | 0.77 | 0.89 | -0.15 | 0.22 |
|  | SAT score | 26.5 (8.52) | 28.6 (9.99) | 2.07 (5.03) | 0.83 | 0.73 | 0.90 | -7.99 | 12.1 |
| Colour Mix Level 3 | Total correct | 39.1 (8.22) | 41.4 (7.50) | 2.23 (3.62) | 0.89 | 0.85 | 0.93 | -5.02 | 9.48 |
|  | Average RT | 1.54 (0.38) | 1.43 (0.36) | -0.11 (0.17) | 0.92 | 0.89 | 0.95 | -0.07 | 0.17 |
|  | SAT score | 28.0 (11.2) | 31.5 (11.6) | 3.56 (4.47) | 0.92 | 0.89 | 0.85 | -5.39 | 12.5 |
| Colour Mix Level 4 | Total correct | 31.3 (9.18) | 33.7 (9.61) | 2.41 (5.15) | 0.85 | 0.78 | 0.90 | -7.89 | 12.7 |
|  | Average RT | 1.95 (0.63) | 1.82 (0.59) | -0.13 (0.36) | 0.87 | 0.81 | 0.91 | -0.37 | 0.23 |
|  | SAT score | 18.8 (9.80) | 21.7 (11.3) | 2.94 (5.31) | 0.87 | 0.81 | 0.91 | -0.92 | 1.55 |
| Think Back Level 1 | Total correct | 31.5 (8.55) | 37.2 (8.24) | 5.65 (5.68) | 0.77 | 0.68 | 0.84 | -5.72 | 17.0 |
|  | Average RT | 1.36 (0.60) | 1.09 (0.47) | -0.27 (0.35) | 0.73 | 0.62 | 0.81 | -0.25 | 0.63 |
|  | SAT score | 28.7 (16.0) | 40.9 (19.9) | 12.2 (13.1) | 0.77 | 0.67 | 0.84 | -1.09 | 3.19 |
| Think Back Level 2 | Total correct | 20.4 (6.38) | 24.1 (6.75) | 3.68 (4.69) | 0.75 | 0.64 | 0.82 | -5.69 | 13.1 |
|  | Average RT | 2.17 (0.89) | 1.75 (0.76) | -0.42 (0.60) | 0.76 | 0.66 | 0.83 | -0.75 | 0.32 |
|  | SAT score | 12.2 (9.13) | 17.4 (11.0) | 5.20 (6.70) | 0.77 | 0.68 | 0.84 | -0.97 | 2.33 |
| Path Finder Level 1 | Completion time | 14.9 (6.21) | 13.2 (4.33) | -1.67 (4.40) | 0.72 | 0.61 | 0.80 | -0.02 | 0.04 |
| Path Finder Level 2 | Completion time | 33.2 (18.2) | 28.6 (15.9) | -4.57 (13.7) | 0.68 | 0.56 | 0.78 | -0.88 | 0.59 |
| Face Match | Total Correct | 27.3 (2.28) | 27.9 (1.86) | 0.62 (1.94) | 0.57 | 0.42 | 0.69 | -3.26 | 4.50 |
|  | Average RT | 1.74 (0.38) | 1.64 (0.39) | -0.11 (0.27) | 0.75 | 0.65 | 0.83 | -0.65 | 0.43 |
|  | SAT score | 16.7 (4.79) | 18.3 (5.54) | 1.68 (3.60) | 0.75 | 0.65 | 0.83 | -0.66 | 1.05 |
| Mind Reading | Total Correct | 11.5 (3.64) | 13.0 (2.96) | 1.47 (2.73) | 0.66 | 0.54 | 0.76 | -3.98 | 6.92 |
|  | Average RT | 4.96 (1.43) | 4.19 (1.06) | -0.77 (1.07) | 0.73 | 0.62 | 0.81 | -0.55 | 0.23 |
|  | SAT score | 2.67 (1.35) | 3.40 (1.38) | 0.74 (0.86) | 0.75 | 0.65 | 0.83 | -0.34 | 0.81 |
| Picture Pair | Total Correct | 22.2 (2.61) | 22.9 (1.83) | 0.73 (1.93) | 0.63 | 0.50 | 0.74 | -3.13 | 4.59 |
|  | Average RT | 3.80 (1.27) | 2.93 (0.99) | -0.86 (0.75) | 0.84 | 0.77 | 0.89 | -0.62 | 0.11 |
|  | SAT score | 6.64 (2.61) | 8.68 (2.77) | 2.04 (1.40) | 0.85 | 0.79 | 0.90 | -0.16 | 0.92 |
| Line Judge | Total Correct | 10.3 (2.32) | 10.3 (2.62) | 0.01. (1.69) | 0.77 | 0.67 | 0.84 | -3.37 | 3.39 |
|  | Average RT | 4.59 (1.33) | 3.99 (1.16) | -0.60 (0.77) | 0.83 | 0.76 | 0.89 | -0.46 | 0.18 |
|  | SAT score | 2.45 (0.96) | 2.81 (1.13) | 0.36 (0.65) | 0.82 | 0.74 | 0.87 | -0.30 | 0.50 |
| Balloon Fair | Total earned | 1145.9 (456.1) | 1369.8 (516.8) | 223.9 (399.3) | 0.68 | 0.56 | 0.77 | -7.80 | 14.2 |
| Swipe Out | Flanker effect (ms) | 265.0 (258.1) | 204.1 (168.5) | -60.9 (230.0) | 0.44 | 0.27 | 0.59 | -520.9 | 399.1 |
|  | Average RT | 1.28 (0.32) | 1.13 (0.27) | -0.15 (0.17) | 0.89 | 0.85 | 0.93 | -0.08 | 0.28 |
| Card Sort | Number of categories completed | 2.37 (1.34) | 2.86 (1.18) | 0.49 (1.03) | 0.67 | 0.54 | 0.76 | -1.57 | 2.55 |
| Time Tap - Paced | Clock variance (ms2) | -1190.2 (42156.7) | -4916.3 (27496.2) | -3726.0 (53158.8) | -0.12 | -0.31 | 0.09 | -1129.0 | 118705.0 |
|  | Absolute drift (ms) | 95.7 (92.3) | 86.8 (89.3) | -8.90 (131.9) | 0.12 | -0.09 | 0.31 | -3.03 | 2.98 |
| Time Tap - Self-paced | Clock variance (ms2) | 6300.1 (21121.8) | 5617.6 (18194.75) | -682.5 (28837.1) | -0.07 | -0.27 | 0.14 | -5.80 | 5.77 |
|  | Absolute drift (ms) | 112.5 (97.9) | 128.9 (101.9) | 16.4 (139.9) | 0.02 | -0.18 | 0.22 | -12.0 | 13.9 |

#### User Experience questionnaire

No significant differences in responses were found between younger and older participants on any of the items. Using a five-point scale all participants (100%) reported (Neutral or above) that the test example videos were helpful, they found the tests interesting, and they enjoyed completing the assessment. Additionally, almost all participants (Neutral or above) agreed that the task instructions were easy to understand (96.4%), the tests were not boring or repetitive (94.5%), the assessment was not difficult to complete from home (98.2%), and the iPad was not difficult to use (98.2%) (Figure 4).

## Discussion

The Ignite app was tested in a population of 2,004 healthy controls, generating the largest normative dataset to date for a computerised cognitive assessment of FTD. The results from Study 1, revealed the Ignite tests capture well-established trajectories of age-related cognitive decline seen in traditional pen and paper versions of tasks on measures of processing speed (24–26), executive function (27,28), social cognition (29–32), semantic knowledge (33,34), and visuospatial skills (35). Notably, there was a strong decline in performance with age on the Path Finder and Colour Mix tasks, corroborating studies of traditional versions of these tests that show increasing completion times with age on the Trail Making Test (24,25), and the Stroop task, respectively (36). Only performance on the Sum Up task did not decline with age. However, mental arithmetic is a complex task that relies on crystallised intelligence, which is known to be more protected from age-related decline (37). Several significant differences in sex were observed for the Ignite tests, with female participants scoring higher on tasks assessing social cognition, replicating well established findings that sex influences performance on emotion processing (38), and empathy tests (39). Female participants also performed higher on Colour Mix Levels 1-3, whilst male subjects performed better on Think Back Level 2, Sum Up, Line Judge, and Balloon Fair. This corroborates the literature describing sex differences in cognitive function that suggest females score higher on tasks assessing cognitive flexibility and inhibitory control, whilst men tend to perform better on working memory, visuospatial processing and decision-making tasks (40–44). Several significant associations with education were observed for the Ignite tests, however, this was likely driven by the large sample size as the associations were small in magnitude. A regression-based normative calculator was subsequently developed that will be useful for assessing cognitive performance at the individual level in future studies. Utilising adjusted *Z*-scores to assess performance on an individual basis could increase the sensitivity of the Ignite tests in detecting subtle deficits in presymptomatic FTD mutation carriers.

The Ignite tests were found to load onto factors associated with expected constructs, suggesting the tests are capturing the hypothesised cognitive domains. Tests of social cognition (Face Match and Mind Reading) and semantic knowledge (Picture Pair) unexpectedly loaded together. However, one theoretical perspective has grouped these processes together within the controlled semantic cognition (CSC) framework (45,46) which represents a conceptual knowledge base of the meaning of words, objects, and people, including person identification, empathy, and emotion recognition (46,47). Additionally, the loading of the Line Judge and Swipe Out task together, could be explained by the likelihood that the tests tap into multiple processes including visual attention and inhibitory control.

The results from Study 2 demonstrated that most of the Ignite tests exhibit good concordance with their corresponding pen and paper counterparts, supporting concurrent validity. Additionally, Ignite tests without direct pen and paper comparisons correlated with other tests that measure the same hypothesised cognitive domains. The only tests that were not significantly associated with traditional measures were the Face Match, Card Sort, and Time Tap tasks. The lack of association between Face Match/Ekman Faces Test and Card Sort/Wisconsin Card Sorting Task can likely be explained by the simplicity of the pen and paper versions of these tasks and the resulting ceiling effects observed. Ceiling effects restrict the range of scores and result in low correlation coefficients. Furthermore, a high degree of variability was observed in the Time Tap data in both outcome measures, suggesting perhaps that cognitive timing is not a stable trait and could be highly influenced by other factors. This test also displayed poor test-retest reliability.

The Ignite tests displayed moderate to excellent test-retest reliability overall. Good test-retest coefficients >0.75 (23) were obtained for all levels of the Colour Mix and Think Back tasks, as well as Sum Up, Line Judge, Face Match average RT and SAT scores, Picture Pair average RT scores, and Swipe Out average RT scores. Thus, findings are also consistent with previous studies demonstrating good reliability in pen and paper versions of these tasks (48–56). Ignite reliability estimates for Think Back levels of working memory were higher than previously reported from 1-back (56,57) and 2-back tasks (56). Consistent with prior research studies using pen and paper (58) and computerised Trail Making Tests (17,59), the Ignite Path Finder levels displayed moderate reliability.

The results of the Ignite User Experience questionnaire demonstrate that healthy controls rate the app favourably overall. The majority of participants reported that the image quality of the tests was good, the videos were helpful, and the instructions were easy to follow. In addition, healthy controls agreed that the app was easy to complete remotely from home, and the iPad was not difficult to use. Demonstrating the feasibility of administering novel computerised assessments is equally as important as proving validity and reliability. Therefore, this data indicates the acceptability of the app amongst healthy adults (including those of older ages), and the feasibility of implementing Ignite as a cognitive test in the wider population through remote data collection studies.

There are several limitations to this work. First, participants in Study 1 mainly resided in the United Kingdom, and ethnocultural and socioeconomic status data was not collected. Second, at the time of this study, only participants that spoke English who owned an Apple iPad were able to participate, limiting the generalisability of these findings. The Ignite app has recently been translated into multiple other languages and has since been implemented across the international GENetic FTD initiative (GENFI) study. This will further increase the reach of the assessment to a broader population leading to greater diversity in datasets.

In conclusion, Ignite is appropriate for a broad range of ages and ability levels, and the tests capture cognitive performance reflective of well-established trajectories in normal ageing. In addition, Ignite is reliable upon repeated testing, displays good concordance with gold-standard neuropsychology tests, and the app is well accepted in healthy controls. One exception is the Time Tap task which will be removed from future versions of Ignite based on the results of this initial study. Future work should focus on repeated testing of Ignite through a burst-testing protocol, to investigate the extent of practice effects upon multiple administration. This will be important for clinical trials to define the optimal number of times Ignite should be completed to obtain an accurate depiction of performance. Furthermore, studies should investigate the Ignite app in clinical and preclinical FTD populations to establish if the tests are sensitive to early cognitive impairment. Following further investigation in FTD, the selection of the most sensitive Ignite tests for each genetic group could help to optimise the assessment. It is likely that gene-specific Ignite composite scores would be beneficial to clinical trials in enhancing the sensitivity of detecting cognitive impairment and reducing the required sample sizes.

## Data Availability

All data produced in the present work are contained in the manuscript

https://www.genfi.org/ignite

## Acknowledgements

KM is now an employee of Ionis Pharmaceuticals but was at UCL during her contribution to this work. Ions have neither supported nor taken part in any aspect of this work. We thank Chris Frost and Amy MacDougall for additional statistical advice.

## Data availability

The datasets used and/or analysed during the current study are available from the corresponding author on reasonable request. The normative calculator is available for use on the GENFI website (www.genfi.org/ignite).

## Code availability

The underlying code for this study is not publicly available but may be made available to qualified researchers on reasonable request from the corresponding author.

