## Supplementary material for "Concurrent validity, test-retest reliability, and normative properties of the Ignite app: a cognitive assessment for frontotemporal dementia"

**Convery et al**

**Supplementary Table 1: Outcome measure calculation.**

| *Colour Mix, Think Back, Mind Reading, Face Match, Sum Up, Picture Pair, and Line Judgement* | **Average reaction time (s):** Calculated by averaging the time taken in seconds to respond and select each stimulus across the total number of trials completed in each of these tasks. |
| --- | --- |
|  | **Total correct**: Calculated by totalling the number of correct trials of those completed. |
|  | **SAT score**: the total number of correct items completed divided by the average reaction time in seconds. |
| *Swipe Out* | **Flanker effect (ms):** calculated by subtracting the average reaction time of trials where the flanking arrows are congruent (same direction as central arrow) from the average reaction time of trials where the flanking arrows are incongruent (different direction to central arrow). |
| *Balloon Fair* | **The total amount of money earned:** The total amount of money accrued in the bank at the end of the task, accounting for any incurred losses by burst balloons. |
| *Card Sort* | **The number of correct categories achieved**: (out of a possible total of six) was calculated. A category can only be completed when six consecutive correct rules have been provided. It was not possible to differentiate between perseverative and non-perseverative errors in the Ignite data due to the structure of the raw data output, therefore the number of correct categories achieved was used as an equivalent measure of frontal dysfunction. |
| *Path Finder* | **Time to complete (s):** The total time taken to complete the task. |
| *Time Tap* | The below measures were generated based on the Wing and Kristofferson (47) model of motor response timing, which proposes that the variance in finger tapping is determined by both a “clock” and motor system delay. The clock represents a cognitive internal timekeeper which provides the trigger to initiate a given response or tap, whereas the motor delay is the time between the clock trigger and the response. These systems are thought to be independent processes and vary between responses. Another assumption of the Wing and Kristofferson (47) model is that although each inter-response interval (IRI) is determined by only the clock for that response it is also determined by the motor delay of that response plus the preceding response. This dependency of the motor delay on the previous response imposes a negative correlation between consecutive IRI’s, meaning a shorter IRI is followed by a long IRI or vice versa.  Both the clock and motor delay values are calculated from the estimated variance output from an autoregressive model allowing for the correlation of a variable with its past and future values. The Prais-Winsten method was adopted, which iterates over different values of the autocorrelation coefficient to find one that minimises the sum of squared errors (48). The Prais model was fitted to model inter-response interval for each condition (paced and self-paced) for each participant. To allow for relatively complex drifts in IRI, quadratic terms were fitted for stimulus number for paced and self-paced conditions (49). For both conditions, the first two taps were removed to eliminate atypical early responses and IRI’s that were more than three standard deviations from the predicted value of the model were also excluded since these were thought to be due to measurement error (i.e., missed, or accidental double taps). Clock variance and absolute drift were used as the outcome measures of interest as previous research demonstrated the greatest differences in these measures between bvFTD and controls (13), whereas a measure of motor delay was considered less important for cognitive impairment in FTD.  **Clock variance (C):** Calculated from the Prais model estimation of the residual variance (G0) and covariance between consecutive residuals (G1), as follows: C= G0 + 2*G1  **Absolute drift:** Calculated as the difference in the first and last tap from the modelled IRI regardless of the direction, to provide a measure of the magnitude of the drift. |

**Supplementary Table 2: Ignite Z-score transformations. Transformations were chosen based on which method achieved the best normal distribution for that test (i.e., the highest *p*-value of a Shapiro-Wilk test).**

| **Test** | **Outcome measure** | **Transformation** |
| --- | --- | --- |
| Path Finder Level 1 | Time to complete (s) | Inverse |
| Path Finder Level 2 | Time to complete (s) | Log |
| Colour Mix Level 1 | Average reaction time (s) | Inverse |
| Colour Mix Level 2 | Average reaction time (s) | Inverse |
| Colour Mix Level 3 | Average reaction time (s) | Inverse |
| Colour Mix Level 4 | Average reaction time (s) | Log |
| Colour Mix Level 4 | SAT score | Square root |
| Think Back Level 1 | Average reaction time (s) | 1/Square root |
| Think Back Level 1 | SAT score | Square root |
| Think Back Level 2 | Average reaction time (s) | Log |
| Think Back Level 2 | SAT score | Log |
| Swipe Out | Average reaction time (s) | 1/Square root |
| Mind Reading | Average reaction time (s) | 1/Square root |
| Mind Reading | SAT score | Square root |
| Face Match | Average reaction time (s) | 1/Square root |
| Picture Pair | Average reaction time (s) | Log |
| Line Judge | Average reaction time (s) | Log |
| Line Judge | SAT score | Square root |
| Sum Up | Average reaction time (s) | 1/Square root |
| Sum Up | SAT score | Square root |

**Supplementary Table 3: Ignite normative values. CI=confidence interval, RT=reaction time in seconds, Time=completion time in seconds, SAT= speed accuracy trade-off, F=Female, M=Male. Adjusted means represent observed coefficients output from the regression models.**

|  |  | Age bin | | | | | | Education bin | | | | Sex | |
| --- | --- | --- | --- | --- | --- | --- | --- | --- | --- | --- | --- | --- | --- |
|  |  | 20-29 | 30-39 | 40-49 | 50-59 | 60-69 | 70-80 | 0-9 | 10-12 | 13-16 | >17 | F | M |
| Path Finder Level 1  Time | Adjusted mean | 12.0 | 13.0 | 14.2 | 15.2 | 16.9 | 20.7 | 18.3 | 16.7 | 16.2 | 15.7 | 15.9 | 16.4 |
|  | Lower 95% CI | 11.1 | 12.1 | 13.3 | 14.5 | 16.4 | 20.0 | 16.7 | 16.0 | 15.6 | 15.2 | 15.6 | 15.9 |
|  | Upper 95% CI | 12.9 | 13.9 | 15.0 | 15.8 | 17.4 | 21.3 | 19.9 | 17.4 | 16.7 | 16.1 | 16.3 | 16.9 |
| Path Finder Level 2  Time | Adjusted mean | 24.7 | 27.1 | 29.3 | 33.3 | 36.9 | 47.5 | 37.0 | 38.6 | 34.7 | 34.1 | 34.4 | 36.7 |
|  | Lower 95% CI | 22.3 | 24.7 | 27.1 | 31.6 | 35.5 | 45.8 | 32.9 | 36.7 | 33.3 | 33.0 | 33.5 | 35.3 |
|  | Upper 95% CI | 27.1 | 29.5 | 31.5 | 35.0 | 38.3 | 49.3 | 41.2 | 40.5 | 36.0 | 35.2 | 35.3 | 38.0 |
| Colour Mix Level 1  Average RT | Adjusted mean | 1.01 | 1.09 | 1.18 | 1.26 | 1.32 | 1.48 | 1.28 | 1.29 | 1.26 | 1.26 | 1.25 | 1.30 |
|  | Lower 95% CI | 0.97 | 1.06 | 1.15 | 1.23 | 1.30 | 1.45 | 1.22 | 1.27 | 1.25 | 1.25 | 1.24 | 1.28 |
|  | Upper 95% CI | 1.04 | 1.12 | 1.21 | 1.28 | 1.34 | 1.50 | 1.34 | 1.32 | 1.28 | 1.28 | 1.26 | 1.32 |
| Colour Mix Level 1  Total correct | Adjusted mean | 29.6 | 27.4 | 25.3 | 24.0 | 22.8 | 20.6 | 23.6 | 23.6 | 24.1 | 24.1 | 24.3 | 23.5 |
|  | Lower 95% CI | 29.1 | 26.9 | 24.8 | 23.6 | 22.5 | 20.2 | 22.7 | 23.2 | 23.8 | 23.9 | 24.1 | 23.2 |
|  | Upper 95% CI | 30.1 | 27.9 | 25.8 | 24.3 | 23.1 | 20.9 | 24.5 | 24.0 | 24.4 | 24.4 | 24.5 | 23.8 |
| Colour Mix Level 1  SAT score | Adjusted mean | 30.5 | 26.1 | 22.3 | 20.1 | 18.1 | 15.0 | 19.6 | 19.9 | 20.5 | 20.6 | 20.8 | 19.7 |
|  | Lower 95% CI | 29.6 | 25.3 | 21.5 | 19.4 | 17.6 | 14.3 | 18.1 | 19.2 | 20.1 | 20.2 | 20.4 | 19.2 |
|  | Upper 95% CI | 31.3 | 27.0 | 23.1 | 20.7 | 18.6 | 15.6 | 21.1 | 20.6 | 21.0 | 21.0 | 21.1 | 20.2 |
| Colour Mix Level 2  Average RT | Adjusted mean | 0.93 | 0.97 | 1.03 | 1.10 | 1.15 | 1.23 | 1.12 | 1.13 | 1.09 | 1.10 | 1.12 | 1.10 |
|  | Lower 95% CI | 0.91 | 0.95 | 1.01 | 1.09 | 1.13 | 1.22 | 1.08 | 1.11 | 1.08 | 1.09 | 1.10 | 1.09 |
|  | Upper 95% CI | 0.95 | 1.00 | 1.06 | 1.12 | 1.16 | 1.25 | 1.16 | 1.14 | 1.11 | 1.11 | 1.13 | 1.11 |
| **Colour Mix Level 2**  Total correct | Adjusted mean | 32.1 | 30.5 | 28.9 | 27.2 | 26.1 | 24.3 | 27.0 | 27.0 | 27.5 | 27.4 | 27.1 | 27.5 |
|  | Lower 95% CI | 31.6 | 30.0 | 28.5 | 26.8 | 25.8 | 23.9 | 26.2 | 26.6 | 27.2 | 27.1 | 26.8 | 27.3 |
|  | Upper 95% CI | 32.6 | 31.0 | 29.4 | 27.5 | 26.4 | 24.7 | 27.9 | 27.3 | 27.8 | 27.6 | 27.3 | 27.7 |
|  |  | **Age bin** | | | | | | **Education bin** | | | | **Sex** | |
|  |  | **20-29** | **30-39** | **40-49** | **50-59** | **60-69** | **70-80** | **0-9** | **10-12** | **13-16** | **>17** | **F** | **M** |
| **Colour Mix Level 2**  SAT score | Adjusted mean | 35.7 | 32.2 | 28.9 | 25.5 | 23.6 | 20.5 | 25.3 | 25.5 | 26.4 | 26.7 | 25.6 | 26.2 |
|  | Lower 95% CI | 34.8 | 31.3 | 28.1 | 24.8 | 23.0 | 19.8 | 23.7 | 24.7 | 25.8 | 25.9 | 25.1 | 25.9 |
|  | Upper 95% CI | 36.7 | 33.2 | 29.8 | 26.2 | 24.1 | 21.2 | 26.9 | 26.2 | 26.9 | 26.9 | 26.2 | 26.6 |
| Colour Mix Level 3  Average RT | Adjusted mean | 1.22 | 1.37 | 1.44 | 1.54 | 1.67 | 1.86 | 1.66 | 1.64 | 1.56 | 1.57 | 1.62 | 1.56 |
|  | Lower 95% CI | 1.18 | 1.32 | 1.40 | 1.51 | 1.65 | 1.83 | 1.58 | 1.60 | 1.53 | 1.55 | 1.59 | 1.55 |
|  | Upper 95% CI | 1.26 | 1.41 | 1.48 | 1.57 | 1.70 | 1.89 | 1.74 | 1.67 | 1.58 | 1.59 | 1.64 | 1.58 |
| **Colour Mix Level 3**  Total correct | Adjusted mean | 45.0 | 42.2 | 39.4 | 37.1 | 34.3 | 29.8 | 34.7 | 34.7 | 36.4 | 37.1 | 35.5 | 36.9 |
|  | Lower 95% CI | 43.8 | 41.0 | 38.2 | 36.2 | 33.6 | 28.9 | 32.5 | 33.8 | 35.8 | 36.6 | 34.8 | 36.4 |
|  | Upper 95% CI | 46.3 | 43.4 | 40.6 | 38.0 | 35.0 | 30.6 | 36.8 | 35.7 | 37.1 | 37.7 | 36.2 | 37.3 |
| **Colour Mix Level 3**  SAT score | Adjusted mean | 43.3 | 40.8 | 37.1 | 34.8 | 32.0 | 26.4 | 32.1 | 31.5 | 33.6 | 34.4 | 32.9 | 34.6 |
|  | Lower 95% CI | 41.1 | 38.6 | 35.1 | 33.2 | 30.7 | 24.8 | 28.3 | 29.9 | 32.4 | 34.4 | 31.7 | 33.8 |
|  | Upper 95% CI | 45.5 | 43.0 | 39.1 | 36.4 | 33.3 | 27.9 | 35.9 | 33.2 | 34.8 | 36.4 | 34.1 | 35.5 |
| Colour Mix Level 4  Average RT | Adjusted mean | 1.46 | 1.71 | 1.86 | 2.05 | 2.30 | 2.74 | 2.18 | 2.32 | 2.08 | 2.12 | 2.19 | 2.12 |
|  | Lower 95% CI | 1.36 | 1.61 | 1.77 | 1.98 | 2.24 | 2.67 | 2.01 | 2.24 | 2.02 | 2.08 | 2.13 | 2.08 |
|  | Upper 95% CI | 1.56 | 1.81 | 1.95 | 2.12 | 2.36 | 2.81 | 2.35 | 2.39 | 2.13 | 2.17 | 2.24 | 2.16 |
| **Colour Mix Level 4**  Total correct | Adjusted mean | 39.3 | 34.8 | 31.8 | 28.8 | 25.6 | 21.0 | 26.5 | 26.4 | 28.7 | 28.9 | 28.0 | 28.5 |
|  | Lower 95% CI | 38.2 | 33.7 | 30.8 | 28.0 | 25.0 | 20.2 | 24.6 | 25.5 | 28.1 | 28.4 | 27.4 | 28.1 |
|  | Upper 95% CI | 40.4 | 35.9 | 32.8 | 29.6 | 26.3 | 21.8 | 28.4 | 27.2 | 29.3 | 29.4 | 28.6 | 29.0 |
| **Colour Mix Level 4**  SAT score | Adjusted mean | 28.6 | 22.3 | 18.9 | 15.7 | 12.8 | 9.1 | 14.0 | 14.7 | 16.4 | 16.1 | 15.7 | 16.0 |
|  | Lower 95% CI | 27.6 | 21.3 | 18.0 | 15.0 | 12.2 | 8.3 | 12.2 | 13.9 | 15.8 | 15.7 | 15.1 | 15.6 |
|  | Upper 95% CI | 29.7 | 23.4 | 19.9 | 16.5 | 13.4 | 9.8 | 15.8 | 15.5 | 16.9 | 16.6 | 16.3 | 16.4 |
| Think Back Level 1  Average RT | Adjusted mean | 0.93 | 1.07 | 1.17 | 1.27 | 1.36 | 1.59 | 1.27 | 1.36 | 1.30 | 1.26 | 1.31 | 1.28 |
|  | Lower 95% CI | 0.87 | 1.01 | 1.11 | 1.22 | 1.32 | 1.55 | 1.16 | 1.31 | 1.26 | 1.23 | 1.28 | 1.26 |
|  | Upper 95% CI | 1.00 | 1.14 | 1.22 | 1.31 | 1.39 | 1.64 | 1.38 | 1.41 | 1.33 | 1.29 | 1.35 | 1.31 |
|  | | **Age bin** | | | | | | **Education bin** | | | | **Sex** | |
|  |  | **20-29** | **30-39** | **40-49** | **50-59** | **60-69** | **70-80** | **0-9** | **10-12** | **13-16** | **>17** | **F** | **M** |
| Think Back Level 1  Total correct | Adjusted mean | 39.0 | 36.4 | 34.9 | 32.9 | 31.0 | 27.8 | 32.3 | 31.3 | 32.4 | 33.2 | 32.4 | 32.6 |
|  | Lower 95% CI | 38.0 | 35.4 | 34.0 | 32.2 | 30.4 | 27.1 | 30.6 | 30.6 | 31.8 | 32.7 | 31.9 | 32.3 |
|  | Upper 95% CI | 40.0 | 37.4 | 35.8 | 33.6 | 31.6 | 28.5 | 33.9 | 32.1 | 32.9 | 33.6 | 33.0 | 33.0 |
| Think Back Level 1  SAT score | Adjusted mean | 45.7 | 38.4 | 34.4 | 30.1 | 26.4 | 20.8 | 28.6 | 28.1 | 30.0 | 31.1 | 30.3 | 30.1 |
|  | Lower 95% CI | 43.7 | 36.4 | 32.5 | 28.7 | 25.3 | 19.3 | 25.2 | 26.5 | 28.9 | 30.2 | 29.2 | 29.3 |
|  | Upper 95% CI | 47.7 | 40.4 | 36.2 | 31.5 | 27.6 | 22.2 | 32.0 | 29.6 | 31.1 | 32.0 | 31.5 | 30.8 |
| Think Back Level 2  Average RT | Adjusted mean | 1.51 | 1.78 | 1.88 | 2.04 | 2.03 | 2.32 | 2.04 | 2.01 | 1.99 | 1.98 | 1.91 | 2.03 |
|  | Lower 95% CI | 1.39 | 1.66 | 1.77 | 1.96 | 1.96 | 2.24 | 1.83 | 1.92 | 1.92 | 1.93 | 1.85 | 1.98 |
|  | Upper 95% CI | 1.63 | 1.89 | 1.98 | 2.12 | 2.09 | 2.40 | 2.24 | 2.10 | 2.05 | 2.04 | 1.98 | 2.07 |
| Think Back Level 2  Total correct | Adjusted mean | 25.9 | 23.2 | 22.4 | 21.0 | 20.7 | 18.8 | 20.5 | 21.0 | 21.2 | 21.6 | 21.9 | 21.1 |
|  | Lower 95% CI | 25.0 | 22.3 | 21.5 | 20.3 | 20.2 | 18.1 | 18.9 | 20.3 | 20.7 | 21.2 | 21.4 | 20.7 |
|  | Upper 95% CI | 26.8 | 24.1 | 23.2 | 21.6 | 21.2 | 19.4 | 22.0 | 21.7 | 21.7 | 22.0 | 22.4 | 21.4 |
| Think Back Level 2  SAT score | Adjusted mean | 21.1 | 17.1 | 15.4 | 13.2 | 12.9 | 10.8 | 13.7 | 14.4 | 14.0 | 14.0 | 15.3 | 13.5 |
|  | Lower 95% CI | 19.7 | 15.8 | 14.1 | 12.2 | 12.1 | 9.8 | 11.3 | 13.4 | 13.3 | 13.4 | 14.6 | 13.0 |
|  | Upper 95% CI | 22.5 | 18.5 | 16.7 | 14.2 | 13.7 | 11.8 | 16.1 | 15.5 | 14.8 | 14.7 | 16.1 | 14.0 |
| Balloon Fair  Total money | Adjusted mean | 1354 | 1306 | 1284 | 1150 | 1112 | 908 | 1124 | 1103 | 1140 | 1162 | 1202 | 1116 |
|  | Lower 95% CI | 1297 | 1249 | 1231 | 1109 | 1078 | 867 | 1025 | 1059 | 1108 | 1136 | 1169 | 1094 |
|  | Upper 95% CI | 1412 | 1363 | 1338 | 1191 | 1146 | 949 | 1224 | 1148 | 1171 | 1189 | 1234 | 1138 |
| Swipe Out  Flanker effect (ms) | Adjusted mean | 216 | 224 | 215 | 241 | 257 | 274 | 240 | 268 | 243 | 238 | 224 | 255 |
|  | Lower 95% CI | 181 | 189 | 182 | 216 | 236 | 249 | 178 | 240 | 224 | 222 | 204 | 241 |
|  | Upper 95% CI | 252 | 259 | 248 | 266 | 278 | 300 | 302 | 295 | 263 | 255 | 243 | 269 |
| Swipe Out  Average RT | Adjusted mean | 0.99 | 1.09 | 1.15 | 1.25 | 1.36 | 1.57 | 1.33 | 1.34 | 1.29 | 1.27 | 1.27 | 1.30 |
|  | Lower 95% CI | 0.96 | 1.06 | 1.12 | 1.23 | 1.34 | 1.55 | 1.27 | 1.32 | 1.27 | 1.25 | 1.25 | 1.29 |
|  | Upper 95% CI | 1.03 | 1.12 | 1.18 | 1.27 | 1.38 | 1.60 | 1.39 | 1.37 | 1.31 | 1.29 | 1.29 | 1.32 |
|  | | **Age bin** | | | | | | **Education bin** | | | | **Sex** | |
|  |  | **20-29** | **30-39** | **40-49** | **50-59** | **60-69** | **70-80** | **0-9** | **10-12** | **13-16** | **>17** | **F** | **M** |
| Card Sort  Correct categories | Adjusted mean | 3.18 | 2.75 | 2.30 | 1.93 | 1.65 | 1.31 | 1.79 | 1.76 | 1.98 | 2.07 | 2.01 | 1.96 |
|  | Lower 95% CI | 3.02 | 2.59 | 2.16 | 1.82 | 1.56 | 1.19 | 1.52 | 1.64 | 1.90 | 1.99 | 1.92 | 1.90 |
|  | Upper 95% CI | 3.34 | 2.91 | 2.45 | 2.04 | 1.74 | 1.42 | 2.07 | 1.88 | 2.07 | 2.14 | 2.09 | 2.02 |
| Face Match  Average RT | Adjusted mean | 1.38 | 1.55 | 1.66 | 1.84 | 1.95 | 2.24 | 1.92 | 1.84 | 1.85 | 1.85 | 1.91 | 1.82 |
|  | Lower 95% CI | 1.30 | 1.48 | 1.59 | 1.79 | 1.90 | 2.19 | 1.79 | 1.78 | 1.81 | 1.82 | 1.87 | 1.79 |
|  | Upper 95% CI | 1.45 | 1.63 | 1.73 | 1.89 | 1.99 | 2.29 | 2.05 | 1.89 | 1.90 | 1.88 | 1.95 | 1.85 |
| Face Match  Total correct | Adjusted mean | 28.7 | 28.1 | 27.8 | 27.1 | 26.7 | 25.0 | 26.6 | 26.9 | 26.9 | 27.0 | 26.4 | 27.2 |
|  | Lower 95% CI | 28.4 | 27.8 | 27.5 | 26.9 | 26.5 | 24.7 | 26.0 | 26.6 | 26.8 | 26.8 | 26.2 | 27.1 |
|  | Upper 95% CI | 29.1 | 28.5 | 28.2 | 27.4 | 26.9 | 25.2 | 27.2 | 27.1 | 27.1 | 27.2 | 26.6 | 27.3 |
| Face Match  SAT score | Adjusted mean | 22.0 | 19.4 | 18.0 | 15.9 | 14.9 | 12.4 | 15.7 | 16.4 | 16.2 | 16.0 | 15.5 | 16.4 |
|  | Lower 95% CI | 21.3 | 18.7 | 17.4 | 15.4 | 14.5 | 11.9 | 14.6 | 15.9 | 15.9 | 15.7 | 15.1 | 16.2 |
|  | Upper 95% CI | 22.7 | 20.1 | 18.6 | 16.4 | 15.3 | 12.9 | 16.9 | 16.9 | 16.6 | 16.3 | 15.9 | 16.7 |
| Mind Reading  Average RT | Adjusted mean | 3.99 | 4.34 | 4.72 | 5.16 | 5.42 | 6.09 | 5.08 | 5.05 | 5.21 | 5.19 | 5.20 | 5.16 |
|  | Lower 95% CI | 3.78 | 4.14 | 4.55 | 5.02 | 5.30 | 5.95 | 4.72 | 4.89 | 5.10 | 5.10 | 5.09 | 5.08 |
|  | Upper 95% CI | 4.19 | 4.55 | 4.93 | 5.31 | 5.54 | 6.24 | 5.43 | 5.21 | 5.32 | 5.29 | 5.31 | 5.24 |
| Mind Reading  Total correct | Adjusted mean | 13.2 | 11.9 | 11.5 | 10.5 | 9.77 | 8.25 | 10.6 | 10.4 | 10.3 | 10.4 | 10.0 | 10.5 |
|  | Lower 95% CI | 12.7 | 11.4 | 11.0 | 10.2 | 9.5 | 7.94 | 9.88 | 10.1 | 10.0 | 10.2 | 9.75 | 10.4 |
|  | Upper 95% CI | 13.6 | 12.3 | 11.9 | 10.9 | 10.0 | 8.56 | 11.4 | 10.8 | 10.5 | 10.6 | 10.2 | 10.7 |
| Mind Reading  SAT score | Adjusted mean | 3.55 | 2.94 | 2.64 | 2.26 | 2.01 | 1.56 | 2.34 | 2.41 | 2.28 | 2.25 | 2.21 | 2.33 |
|  | Lower 95% CI | 3.40 | 2.79 | 2.50 | 2.16 | 1.93 | 1.46 | 2.09 | 2.30 | 2.20 | 2.18 | 2.13 | 2.27 |
|  | Upper 95% CI | 3.69 | 3.08 | 2.77 | 2.36 | 2.10 | 1.66 | 2.59 | 2.52 | 2.36 | 2.32 | 2.29 | 2.38 |
| Picture Pair  Average RT | Adjusted mean | 3.15 | 3.42 | 3.61 | 3.74 | 4.05 | 4.85 | 4.02 | 3.91 | 3.96 | 3.93 | 4.01 | 3.91 |
|  | Lower 95% CI | 3.02 | 3.29 | 3.49 | 3.65 | 3.98 | 4.76 | 3.79 | 3.81 | 3.89 | 3.87 | 3.94 | 3.86 |
|  | Upper 95% CI | 3.28 | 3.55 | 3.72 | 3.84 | 4.13 | 4.94 | 4.24 | 4.01 | 4.03 | 3.99 | 4.08 | 3.96 |
|  | | **Age bin** | | | | | | **Education bin** | | | | **Sex** | |
|  |  | **20-29** | **30-39** | **40-49** | **50-59** | **60-69** | **70-80** | **0-9** | **10-12** | **13-16** | **>17** | **F** | **M** |
| Picture Pair  Total correct | Adjusted mean | 22.0 | 21.9 | 22.4 | 22.6 | 22.3 | 20.6 | 21.1 | 22.0 | 21.9 | 22.2 | 21.7 | 22.1 |
|  | Lower 95% CI | 21.7 | 21.6 | 22.1 | 22.4 | 22.1 | 20.4 | 20.5 | 21.7 | 21.7 | 22.0 | 21.5 | 22.0 |
|  | Upper 95% CI | 22.4 | 22.3 | 22.7 | 22.9 | 22.5 | 20.9 | 21.7 | 22.2 | 22.1 | 22.3 | 21.9 | 22.3 |
| Picture Pair  SAT score | Adjusted mean | 7.32 | 6.80 | 6.61 | 6.31 | 5.84 | 4.64 | 5.65 | 6.10 | 5.95 | 6.09 | 5.96 | 6.07 |
|  | Lower 95% CI | 7.09 | 6.57 | 6.39 | 6.14 | 5.70 | 4.47 | 5.24 | 5.91 | 5.82 | 5.98 | 5.82 | 5.98 |
|  | Upper 95% CI | 7.56 | 7.04 | 6.83 | 6.48 | 5.98 | 4.81 | 6.07 | 6.28 | 6.08 | 6.20 | 6.09 | 6.16 |
| Line Judge  Average RT | Adjusted mean | 3.77 | 4.14 | 4.15 | 4.43 | 4.90 | 5.57 | 4.88 | 4.89 | 4.68 | 4.57 | 4.36 | 4.82 |
|  | Lower 95% CI | 3.58 | 3.95 | 3.98 | 4.30 | 4.79 | 5.44 | 4.56 | 4.74 | 4.58 | 4.48 | 4.26 | 4.75 |
|  | Upper 95% CI | 3.96 | 4.32 | 4.32 | 4.56 | 5.01 | 5.71 | 5.21 | 5.03 | 4.78 | 4.65 | 4.46 | 4.89 |
| Line Judge  Total correct | Adjusted mean | 10.2 | 10.0 | 10.1 | 9.9 | 9.42 | 8.83 | 8.83 | 9.09 | 9.56 | 9.90 | 10.6 | 9.12 |
|  | Lower 95% CI | 9.80 | 9.69 | 9.78 | 9.68 | 9.22 | 8.58 | 8.22 | 8.82 | 9.37 | 9.74 | 10.4 | 8.98 |
|  | Upper 95% CI | 10.51 | 10.38 | 10.43 | 10.18 | 9.62 | 9.08 | 9.44 | 9.36 | 9.75 | 10.1 | 10.8 | 9.25 |
| Line Judge  SAT score | Adjusted mean | 2.93 | 2.64 | 2.61 | 2.40 | 2.12 | 1.78 | 1.98 | 2.16 | 2.29 | 2.37 | 2.70 | 2.10 |
|  | Lower 95% CI | 2.81 | 2.52 | 2.50 | 2.31 | 2.05 | 1.70 | 1.77 | 2.07 | 2.22 | 2.31 | 2.64 | 2.05 |
|  | Upper 95% CI | 3.06 | 2.76 | 2.73 | 2.49 | 2.19 | 1.87 | 2.19 | 2.25 | 2.36 | 2.43 | 2.77 | 2.14 |
| Sum Up  Average RT | Adjusted mean | 3.66 | 3.88 | 3.75 | 3.57 | 3.65 | 3.95 | 4.14 | 3.88 | 3.74 | 3.64 | 3.47 | 3.85 |
|  | Lower 95% CI | 3.50 | 3.72 | 3.60 | 3.46 | 3.56 | 3.84 | 3.86 | 3.75 | 3.65 | 3.57 | 3.38 | 3.79 |
|  | Upper 95% CI | 3.82 | 4.04 | 3.90 | 3.69 | 3.75 | 4.07 | 4.42 | 4.00 | 3.83 | 3.72 | 3.56 | 3.92 |
| Sum Up  Total correct | Adjusted mean | 15.2 | 14.5 | 15.2 | 15.7 | 15.4 | 14.3 | 13.6 | 14.4 | 15.2 | 15.6 | 16.4 | 14.5 |
|  | Lower 95% CI | 14.6 | 13.9 | 14.7 | 15.2 | 15.1 | 13.9 | 12.5 | 13.9 | 14.8 | 15.2 | 16.1 | 14.3 |
|  | Upper 95% CI | 15.8 | 15.2 | 15.8 | 16.1 | 15.8 | 14.8 | 14.7 | 14.9 | 15.5 | 15.8 | 16.8 | 14.7 |
| Sum Up  SAT score | Adjusted mean | 4.89 | 4.45 | 4.79 | 5.01 | 4.89 | 4.24 | 3.80 | 4.34 | 4.77 | 4.91 | 5.53 | 4.34 |
|  | Lower 95% CI | 4.53 | 4.09 | 4.45 | 4.75 | 4.67 | 3.98 | 3.18 | 4.06 | 4.57 | 4.74 | 5.33 | 4.20 |
|  | Upper 95% CI | 5.26 | 4.81 | 5.12 | 5.26 | 5.10 | 4.50 | 4.43 | 4.62 | 4.97 | 5.07 | 5.73 | 4.48 |

**Supplementary Figure 1. Bland-Altman plots constructed for the Ignite outcome measures to demonstrate agreement between scores. The solid line represents the mean difference between scores, and the dashed lines are the 95% upper and lower levels of agreement. The larger circles represent more participants with that value. Figures display: a) normally distributed measures, b) log transformed measures, c) inverse transformed measures and, d) square root transformed measures.**

**a.**

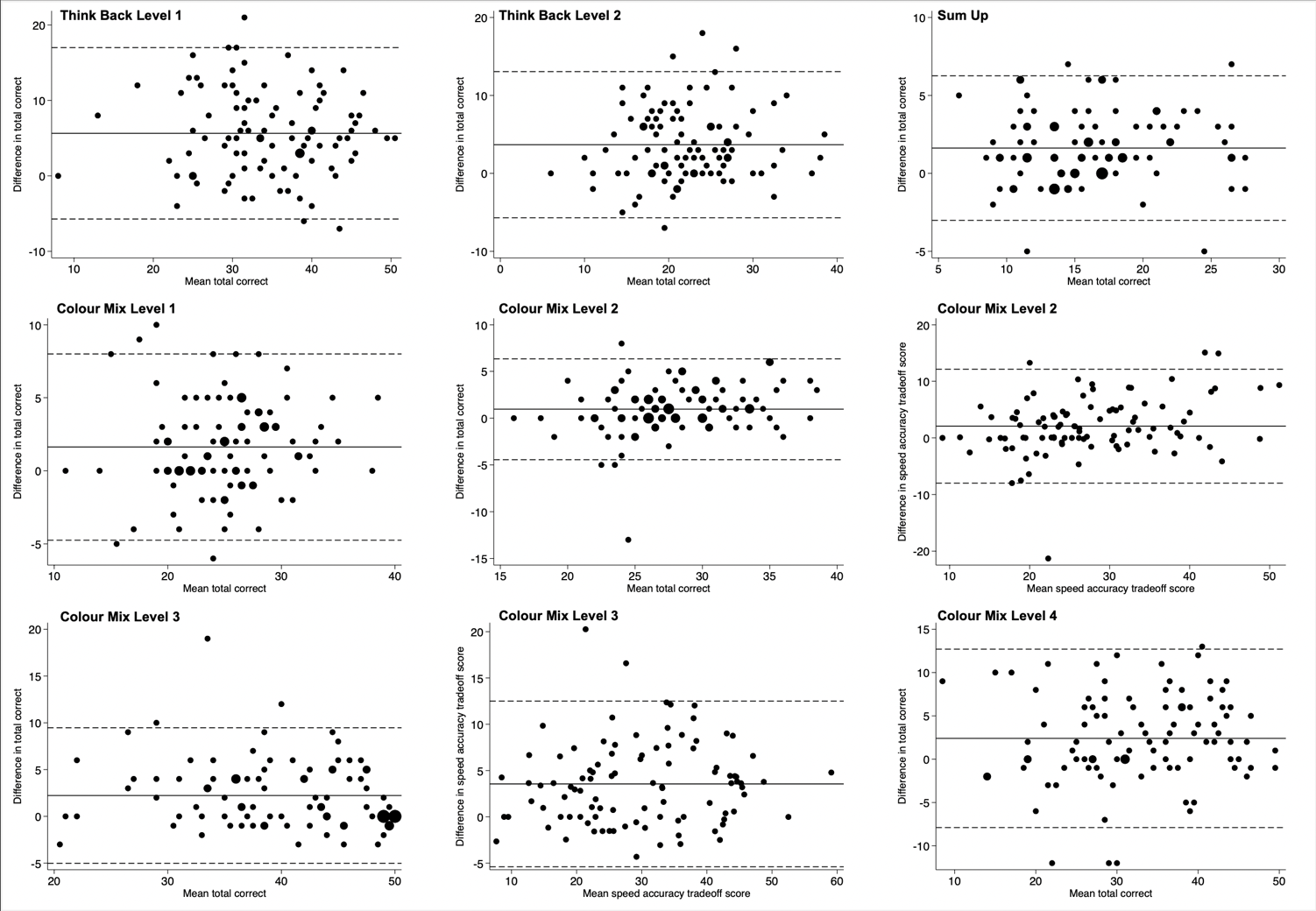

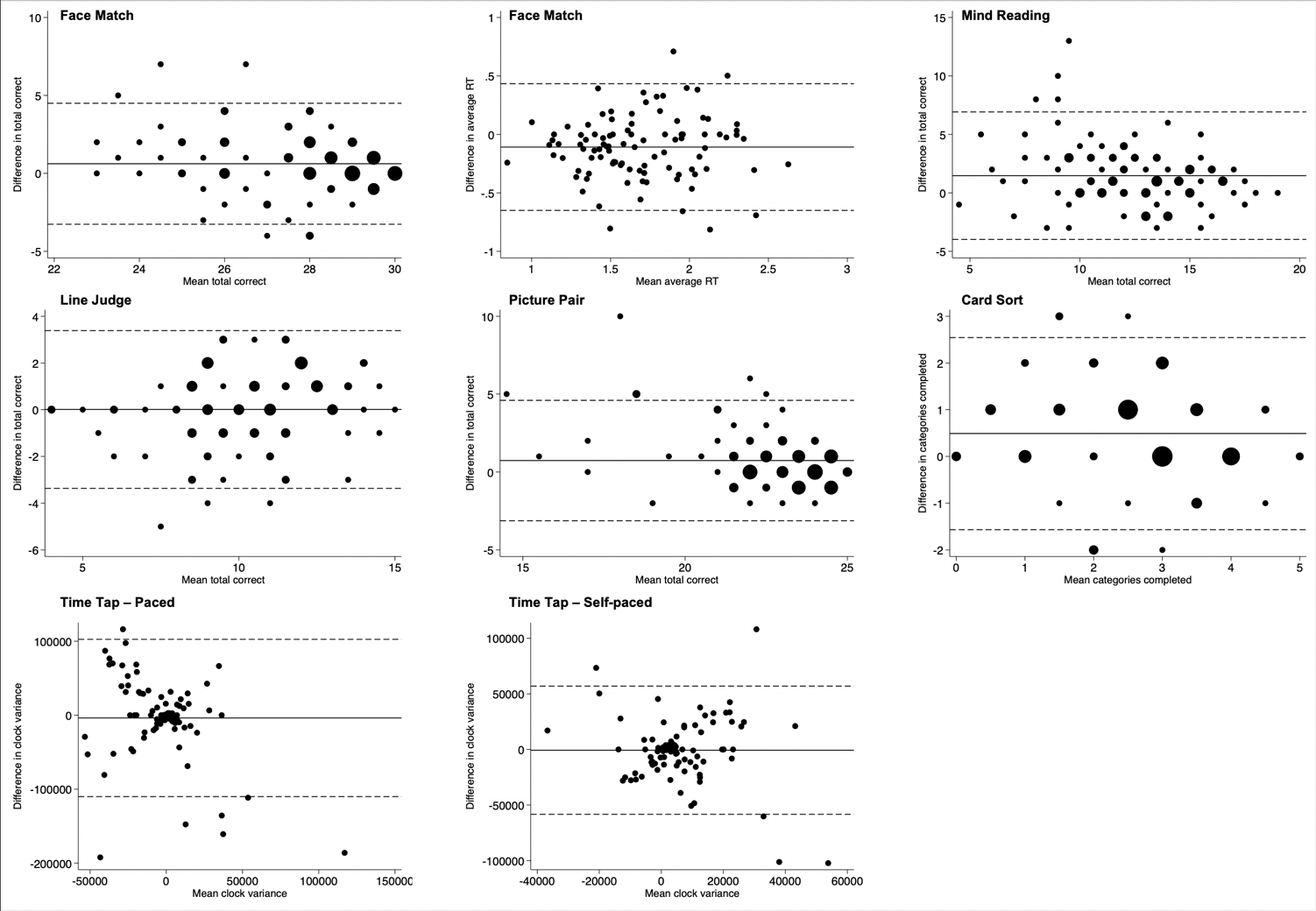

**b.**

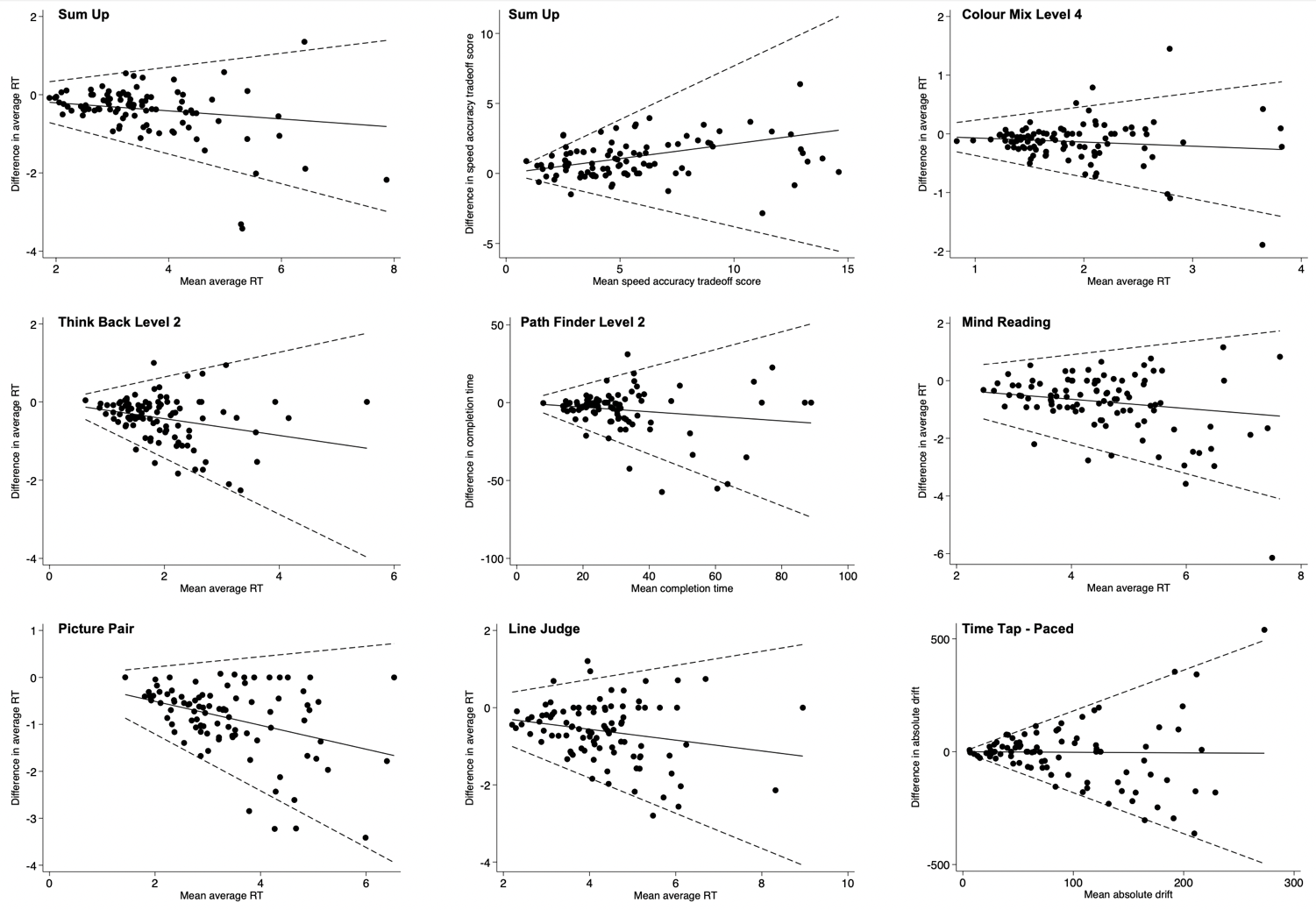

**c.**

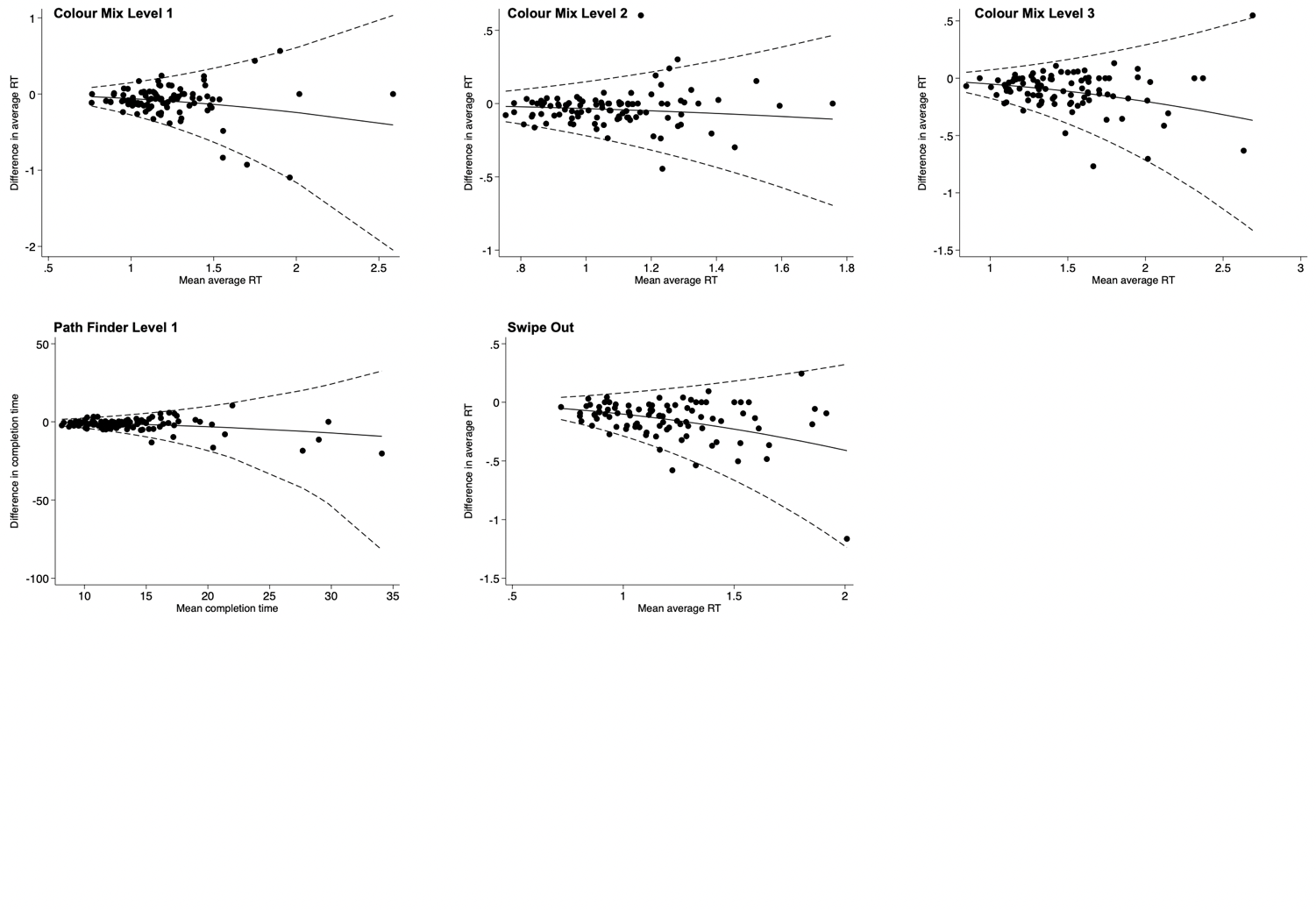

**d.**

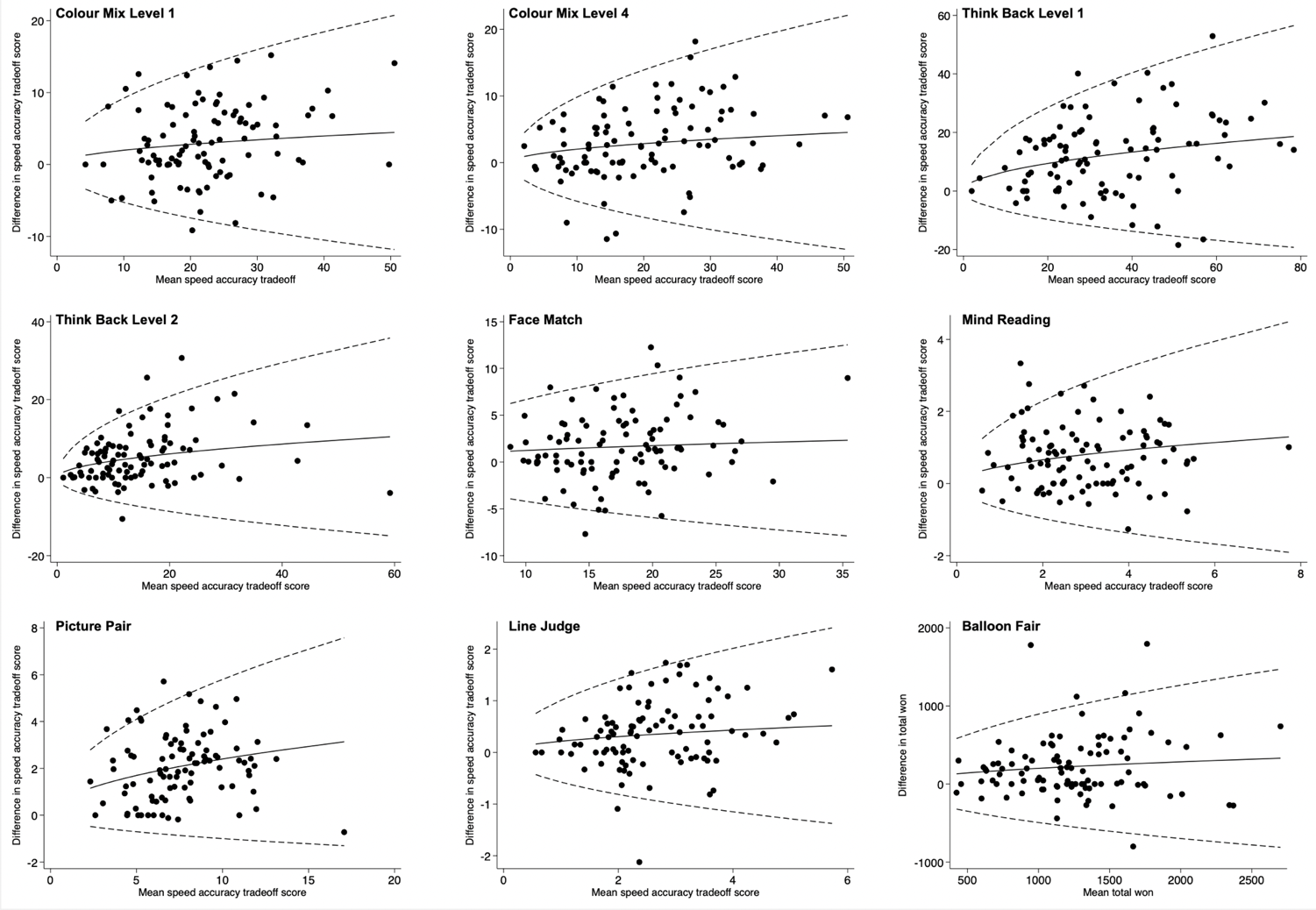
